# Association between male circumcision and human papillomavirus infection in males and females: a systematic review, meta-analysis, and meta-regression

**DOI:** 10.1101/2022.08.08.22278357

**Authors:** Samantha B. Shapiro, Cassandra Laurie, Mariam El-Zein, Eduardo L. Franco

## Abstract

**Background:** Human papillomavirus (HPV) infection is a necessary cause of cervical cancer and is associated with anal, penile, vaginal, and vulvar cancers. Previous studies have suggested a protective effect of male circumcision (MC) on HPV infections in males, and that this protection may be conferred to their female sexual partners. We synthesized the available evidence on the association between MC and HPV infections in males and females.

**Methods:** We performed a systematic review and meta-analysis of the effect of MC on the prevalence, incidence, and clearance of genital HPV infections in heterosexual males and their female sexual partners. We searched multiple databases for studies that assessed MC status and tested for the presence of genital HPV DNA. We used random-effects meta-analysis models to estimate summary measures of effect and 95% confidence intervals (CI) for the prevalence, incidence, and clearance of HPV infections in males and females. We assessed effect modification for prevalence in males using random-effects meta-regression.

**Findings:** We included 32 publications encompassing 25 unique study populations. MC was associated with decreased odds of prevalent HPV infections (odds ratio 0·45, CI 0·34–0·61), a reduced rate of incident HPV infections (incidence rate ratio 0·69, CI 0·57–0·83), and an increased risk of clearing HPV infections (risk ratio 1·44, CI 1·28–1·61) at the glans penis. Effect modification by sampling site was observed for HPV prevalence in males, with greater protection conferred by MC at the glans than the shaft (OR 0·68, 95% CI 0·48–0·98). Females with circumcised sexual partners were at reduced risk for all outcomes.

**Interpretation:** MC protects against various HPV infection outcomes, especially at the glans, and may be a viable prophylactic strategy in regions with a high burden of HPV-associated disease where the HPV vaccine is not commercially available. That the protective effect of MC on HPV infection prevalence varies by penile site has important implications for epidemiologic studies of HPV transmission.

**Funding:** Funded by grant FDN-143347 from the Canadian Institutes of Health Research.

**RESEARCH IN CONTEXT:** *Evidence before this study:* Previous meta-analyses published in 2011, 2012, and 2017 have assessed the impact of MC on genital HPV infections in males, while systematic reviews published in 2017 and 2019 have described the impact of MC on women’s sexual health outcomes. All meta-analyses of males found a protective effect of MC on HPV prevalence, with inconsistent evidence for the association between MC and HPV incidence and clearance. Systematic reviews in females found a protective effect of MC on HPV prevalence.

*Added value of this study:* We identified an additional 12 publications (including one randomized controlled trial) that were not included in the most recently published systematic review and meta-analysis. We found that in males, MC conferred protection against prevalent HPV infections at the glans and shaft of the penis, protected against the acquisition of HPV infections at the glans, and resulted in increased clearance of HPV infections at the glans and shaft. We also found that MC protected females against various HPV infection outcomes. We considered anatomical site in all analyses and explored effect modification using a meta-regression approach. Our meta-analysis also examined the impact of MC on various HPV infection outcomes in females. To our knowledge, the latter two types of analyses had not been done before.

*Implications of all the available evidence:* Countries with a high burden of HPV-associated diseases, or where the HPV vaccine is not commercially available, may wish to consider male circumcision as a preventive strategy. Both males and their female sexual partners may benefit from MC for protection from HPV infections.

## INTRODUCTION

Human papillomavirus (HPV) is the most common sexually transmitted infection worldwide.^1^ Persistent infection with high-risk HPV types (hrHPV) is a necessary cause of cervical cancer and is associated with penile, anal, vaginal, vulvar, and head and neck cancers,^2-4^ while infection with some low-risk HPV types (lrHPV) is associated with genital warts.^1^

Male circumcision (MC) protects against a variety of sexually transmitted infections, including human immunodeficiency virus (HIV), herpes simplex type 2, trichomoniasis, chancroid, and syphilis.^5-7^ Several randomized controlled trials (RCT) evaluating the association between MC and HIV acquisition have also included analyses of HPV as secondary endpoints.^8,9^ Most observational studies of the relationship between MC and HPV infections in males have been cross-sectional in nature, and few have evaluated the risk of HPV infection in female partners of circumcised and uncircumcised males. Previous systematic reviews^10,11^ and meta-analyses^12-14^ found that MC protects against a variety of HPV infection outcomes in males and their female sexual partners. However, gaps in knowledge remain and multiple studies on the topic have been recently published, necessitating an update to the existing literature. In this systematic review, we synthesize the growing evidence suggestive of a protective relationship between MC and HPV infections in males, and the conferred protection to female sexual partners.

## METHODS

### Search strategy and selection criteria

We searched for studies that 1) included participants with no HPV-associated genital lesions, 2) tested for the presence of HPV DNA in genital epithelial cells, 3) assessed the male circumcision status, and 4) assessed the prevalence, incidence, and/or clearance of HPV infections. We included both observational and experimental study designs but excluded case reports and case series. We included studies of both males and females of any age but excluded studies that focused solely on men who have sex with men and people living with HIV from the sample due to HIV’s direct effect on HPV infection risk due to immunosuppression and shared sexual transmission characteristics.^15,16^ Multiple publications from the same study population were eligible for inclusion if they assessed distinct outcomes. We applied no country, date, or language restrictions. We searched the MEDLINE, Embase, Scopus, Cochrane, LILACS, and ProQuest Dissertations & Theses Global databases to identify relevant records published up to 22 June 2022. We also manually searched for potentially eligible studies from previous knowledge syntheses and conference abstracts. The search strategy for each database, developed with input from a university librarian, is included in Supplementary Table 1.

After de-duplicating search results in EndNote, S.S. and C.L. independently screened the abstract of each record to determine relevancy. For papers deemed potentially relevant, we obtained and independently screened the record’s full text. Disagreements at both stages were resolved by consensus.

### Data analysis

S.S. and C.L. performed data extraction using a standardized spreadsheet. Each author extracted data from half of the included records, which was subsequently verified by the alternate author. Extracted data included study characteristics (design, year(s), country(s) and their economic development as defined by the World Bank,^17^ population description, number of visits if longitudinal), exposure and outcome methods (MC assessment method, genital sites sampled, frequency of genital sampling, sampling method, HPV DNA detection and genotyping method, HPV types detected and genotyped), study population results (sample size, sex, age at baseline, HPV prevalence at baseline), and outcome-related data (outcome type, i.e., prevalence, incidence, clearance; HPV risk grouping; number of samples analyzed; number circumcised and uncircumcised; number of prevalent or incident or cleared infections; person-time at risk; effect estimate and 95% confidence interval (95% CI); and covariates adjusted for). Whenever possible, we extracted separate estimates for infection with any HPV type, hrHPV, and lrHPV, as well as separate estimates from samples of different sites of the penis: shaft and/or scrotum only (hereafter referred to as shaft), glans and/or urethra and/or foreskin only (hereafter referred to as glans), and from combinations of shaft sites and glans sites (hereafter referred to as combined site). We extracted the adjusted estimate when available and the crude estimate otherwise. If raw data were presented without effect estimates, we calculated the odds ratio and 95% CI using OpenEpi’s two by two table function.^18^ If effect estimates used circumcised males as the reference category, we took the reciprocal of the estimate and its 95% CI. If relevant data or analyses were mentioned but not quantitatively reported, we contacted the study authors.

We assessed the risk of bias in each study using customized versions of the Newcastle-Ottawa scale for cross-sectional and cohort studies and the Cochrane risk-of-bias tool for randomized trials.^19^ Studies were deemed to have a low risk of bias if they were assigned a score of 7 or greater on the Newcastle-Ottawa scale or a score of low across at least 4 domains using the Cochrane risk-of-bias tool.

We extracted effect estimates for the relationship between MC and HPV infection prevalence, incidence, and clearance for multiple sexes (male and female), HPV risk groupings (any HPV, hrHPV, and lrHPV), and sampling sites (glans, shaft, and combined site). We used the *meta* command in Stata (version 17·0, StataCorp, College Station, Texas) to calculate pooled odds ratios, risk ratios, incidence rate ratios, hazard ratios, and their corresponding 95% CIs using a restricted maximum likelihood model. We assessed study heterogeneity using the I^2^ statistic. We used random effects models for analyses with an I^2^ of greater than or equal to 25% and fixed effects models otherwise. For all analyses, we performed subgroup analyses by sampling site (glans-only vs. shaft-only or combined site) and HPV oncogenicity (hrHPV vs. lrHPV) to assess potential effect modification. We conducted univariate random-effects meta-regression of prevalence studies in males with clustering by study using the *metafor* package^20^ in R (version 4.2.0, R Core Group, Vienna) to explore potential effect modification by study characteristics: year of publication, sites sampled, study country’s economic development, and whether the study controlled for confounding. For studies of prevalence that reported risk or prevalence ratios, we used the raw data to calculate an odds ratio so that the study could be included in the meta-regression.

We performed several sensitivity analyses to assess the robustness of our findings. We repeated our primary analyses of prevalence, incidence, and clearance in males including only studies judged to have a low risk of bias. We additionally repeated our analysis of prevalence in males stratifying by whether studies controlled for confounding, conducted leave-one-out analyses to assess the impact of any one study on the pooled estimate,^21^ and assessed publication bias using a funnel plot and the Egger test.^22^

This study protocol was registered in PROSPERO (registration number CRD42020211591).

## RESULTS

We identified 1,409 potentially eligible records through systematic database searches and 10 through manual searches, of which 624 remained after de-duplication (Figure 1). We excluded 520 records after title and abstract screening, leaving 104 full-text records for assessment. We excluded 18 records for reporting on the same study population as an included record, 25 records that studied participants with HPV-associated lesions, seven records for not sampling a genital site, seven records for missing exposure or outcome data, seven records for not having an outcome of interest, and eight records for failure to obtain needed data that were missing from the authors of the original studies (Supplementary Table 2). In total, we included 32 records in our systematic review and meta-analysis.

**Figure 1:**
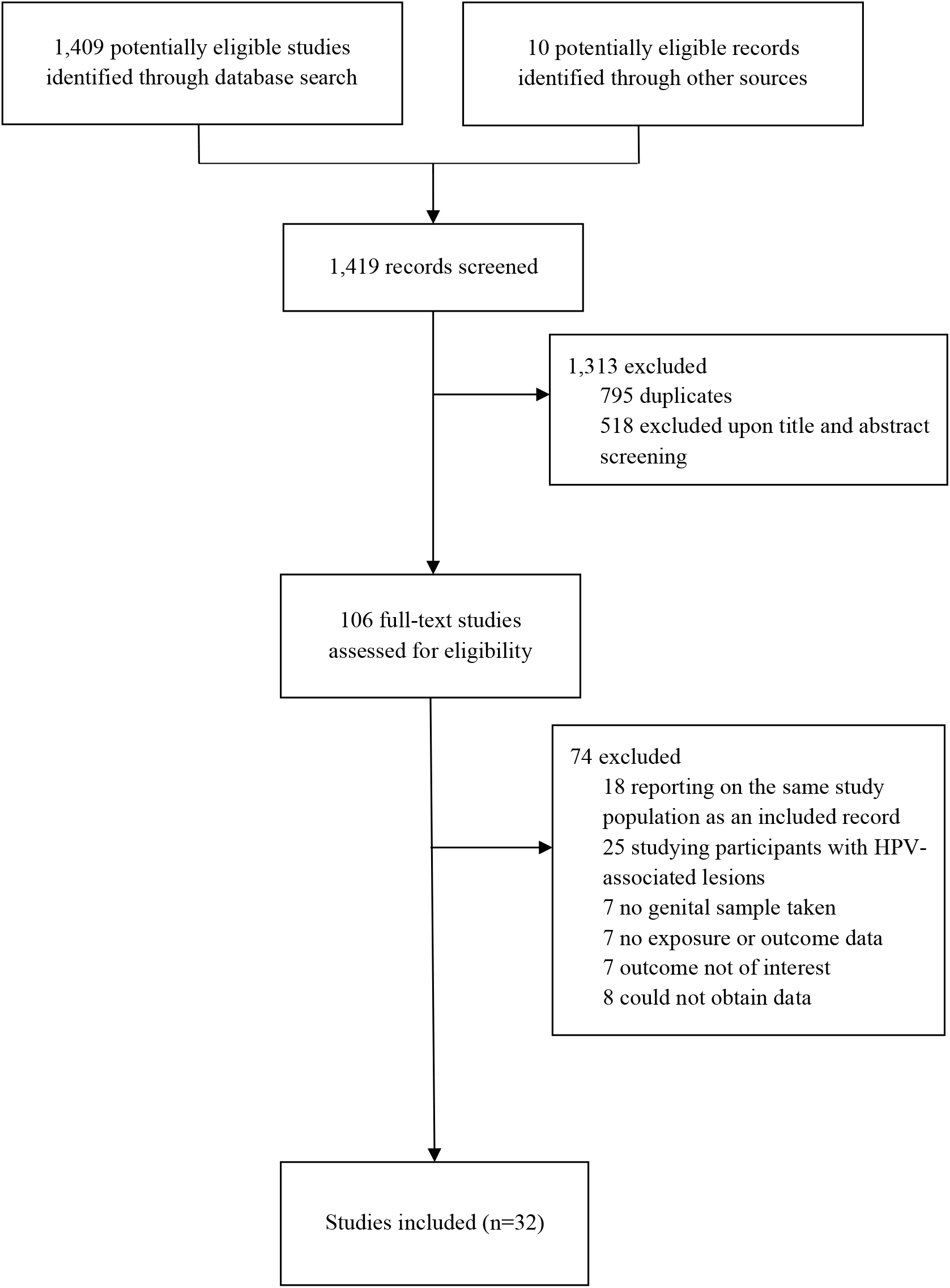
Study selection.

Characteristics of these publications,^6,8,23-52^ which were published between 2002 and 2022, are presented in Table 1. The 32 studies encompassed 25 unique study populations. Of these publications, 17 were cross-sectional studies, ten were cohort studies (of which two were analyzed cross-sectionally), and five were RCTs. Studies were conducted in North America (n=12), South America (n=3), Europe (n=4), Asia (n=1), Africa (n=8), and intercontinentally (n=4). MC status was either self-reported or reported by a partner (n=11), reported by a clinician (n=16), or randomized and verified by a clinician (n=5). All studies assessed the presence of HPV DNA by PCR, 23 of which genotyped for 20 or more HPV types. Samples were taken via swab (n=18), textured paper and swab (n=6), brush (n=5), and brush and swab (n=2). Samples in males were taken from multiple sites, including the urethra (n=5), foreskin (n=15), glans and/or corona (n=27), shaft (n=19), scrotum (n=15), and perianal area (n=4) whereas samples in females were taken from the cervix and vagina (n=5). The PCR primer sets used for HPV DNA typing were PGMY09/11 (n=13), MY09/11 (n=8), GP5+/6+ (n=4), SPF10 (n=3), CpI/CpIIG (n=1), and type-specific and assay-specific primers (n=3). HPV prevalence among all participants at baseline ranged from 8·7% to 69·8%.

**Table 1:** Characteristics of included records according to study design.

| First author (year) | Country(ies), years conducted | World Bank economic classification | Study population | Number enrolled | Circumcision assessment | Sites sampled | HPV DNA genotyping method |
| --- | --- | --- | --- | --- | --- | --- | --- |
| *Randomized controlled trials* |  |  |  |  |  |  |  |
| Gray (2010) | Uganda, 2003–2006 | Low-income country | Males enrolled in the Rakai-1 trial | 840 | Randomized and verified by a clinician | Glans | MY09/11 PCR |
| Smith (2021) | Kenya, 2002–2005 | Middle-income country | Males enrolled in the Kisumu circumcision trial | 2,193 | Randomized and verified by a clinician | Inner foreskin, glans, outer foreskin, shaft | GP5+/6+ PCR |
| Tobian (2009) | Uganda, 2003–2007 | Low-income country | Males enrolled in the Rakai-1 and Rakai-2 trials | 3,393 | Randomized and verified by a clinician | Foreskin, glans | PGMY09/11 PCR |
| Tobian (2012) | Uganda, 2002–2009 | Low-income country | HIV-positive and negative males enrolled in the Rakai-1 and Rakai-2 trials | 776<sup>a</sup> | Randomized and verified by a clinician | Glans | PGMY09/11 PCR |
| Wawer (2011) | Uganda, 2003–2007 | Low-income country | Female partners of males enrolled in the Rakai-1 and Rakai-2 trials | 1,245 | Randomized and verified by a clinician | Vagina | MY09/11 PCR |
| *Cohort studies* |  |  |  |  |  |  |  |
| Albero (2013)<sup>b</sup> | Brazil, Mexico, United States, 2005–2009 | Predominantly high-income countries | Males from the general population, universities, and organized healthcare systems | 3,969 | Clinical exam | Foreskin, glans, shaft, scrotum | PGMY09/11 PCR |
| Albero (2014) | Brazil, Mexico, United States, 2005–2009 | Predominantly high-income countries | Males from the general population, universities, and organized healthcare systems | 4,003 | Clinical exam | Foreskin, glans, shaft, scrotum | PGMY09/11 PCR |
| Hernandez (2008)<sup>b</sup> | United States, 2004–2006 | High-income country | Male university students in Hawaii | 379 | Clinical exam | Foreskin, glans, shaft, scrotum | PGMY09/11 PCR |
| Hernandez (2010) | United States, 2004–2006 | High-income country | Male university students in Hawaii | 357 | Clinical exam | Foreskin, glans, shaft, scrotum | PGMY09/11 PCR |
| Lajous (2005) | Mexico, 2002–2005 | Middle-income country | Healthy military males | 1,030 | Self-report | Urethra, glans, shaft, scrotum | MY09/11 PCR |
| Lu (2009) | United States, 2003–2006 | High-income country | Males from the general population | 285 | Self-report | Foreskin, glans, shaft, scrotum | PGMY09/11 PCR |
| Nielson (2009) | United States, 2002–2005 | High-income country | Males from the general population | 463 | Self-report | Foreskin, urethra, glans, shaft, scrotum, perianal area, anus | PGMY09/11 PCR |
| Partridge (2007) | United States, 2003–2006 | High-income country | Male university students in Washington | 240 | Clinical exam | Foreskin, urethra, glans, shaft, scrotum | PGMY09/11 PCR |
| Shapiro (2022) | Canada, 2005–2011 | High-income country | Female university students in Montreal and their male sexual partners | 826 | Clinical exam | Foreskin, glans, shaft | PGMY09/11 PCR |
| VanBuskirk (2011) | United States, 2003–2009 | High-income country | Male university students in Washington | 477 | Clinical exam | Foreskin, glans, shaft, scrotum | MY09/11 PCR |
| *Cross-sectional studies* |  |  |  |  |  |  |  |
| Baldwin (2004) | United States, 2000–2001 | High-income country | Males attending an STI clinic | 393 | Clinical exam | Glans | PGMY09/11 PCR |
| Bleeker (2005) | Netherlands, 1995–2002 | High-income country | Males from a non-STI dermatology clinic and male partners of females with CIN | 356 | Clinical exam | Foreskin, glans | GP5+/6+ PCR |
| Castellsagué (2002) | Spain, Colombia, Brazil, Thailand, Philippines, 1985–1993 | Predominantly middle-income countries | Male partners of case females with cervical cancer and healthy control females | 1,913 | Clinical exam | Urethra, glans | MY09/11 PCR |
| Contreras (2008) | Mexico, 2005–2006 | Middle-income country | Females with rheumatoid arthritis | 61 | Self-report | Cervix | CpI/CpIIg PCR |
| Da Rocha (2015) | Brazil, 2011–2013 | Middle-income country | Males from an STI clinic, a dermatology clinic, a university, and a factory | 261 | Self-report | Glans | MY09/11 PCR |
| Hebnes (2021) | Denmark, 2006–2007 | High-income country | Military males | 2,460 | Clinical exam | Preputial cavity, glans, shaft, scrotum, perineum | SPF10 PCR |
| Mbulawa (2009) | South Africa, NR | Middle-income country | Sexually active Black heterosexual couples | 254<sup>a</sup> | Self-report | Foreskin, glans, shaft | PGMY09/11 PCR |
| Obiri-Yeboah (2017) | Ghana, NR | Middle-income country | Females attending an HIV or medical outpatient clinic | 170<sup>a</sup> | Partner report | Cervix | RT-PCR with type-specific primers |
| Ogilvie (2009) | Canada, NR | High-income country | Heterosexual males attending an STI clinic | 262 | Clinical exam | Foreskin, glans, shaft, scrotum | Amplicor® primer PCR |
| Olesen (2019) | Tanzania, 2009 | Middle-income country | Males from urban and rural areas | 1,902<sup>a</sup> | Clinical exam | Foreskin, glans, shaft | PGMY09/11 PCR |
| Rocha (2012) | Brazil, NR | Middle-income country | Heterosexual couples in which the female HPV-related cervical lesions | 43 | Clinical exam | Foreskin, glans | GP5+/6+ PCR |
| Rombaldi (2006) | Brazil, 2003–2004 | Middle-income country | Male sexual partners of females with CIN | 99 | Self-report | Foreskin, urethra, glans, shaft | MY09/11 PCR |
| Roura (2012) | Spain, 2007–2008 | High-income country | Females attending cervical cancer screening | 3,261 | Partner report | Cervix | SPF10 PCR |
| Shin (2004) | South Korea, 2002 | High-income country | Male university students | 381 | Self-report | Urethra, glans, shaft, scrotum | SPF10 PCR |
| Svare (2002) | Denmark, 1993 | High-income country | Males attending an STI clinic | 198 | Self-report | Glans, shaft, scrotum, perianal area | GP5+/6+ PCR |
| Vaccarella (2006) | Mexico, 2003–2004 | Middle-income country | Males requesting a vasectomy | 779 | Clinical exam | Glans, shaft, scrotum | MY09/11 PCR |
| Vardas (2011) | 18 countries in Africa, Asia-Pacific, Europe, Latin America, and North America, NR | Predominantly middle-income countries | Heterosexual males with 1–5 female lifetime sexual partners | 3,463 | Clinical exam | Penis (specific sites NR), scrotum, perianal area | RT-PCR with type-specific primers |

Abbreviations: NR, not reported; STI: sexually transmitted infection
<sup>a</sup> Only HIV-negative males were included
<sup>b</sup> Cohort study analyzed cross-sectionally

A total of 21 studies reported estimates for the association between MC and prevalent HPV infections in males (Supplementary Table 3). Sample sizes ranged from 37 to 3,969. MC was associated with significantly decreased odds of prevalent HPV infections at both the glans (OR 0·45, 95% CI 0·34–0·61, I^2^=0·0%) and the shaft or combined sites (OR 0·66, 95% CI 0·50–0·87, I^2^=67·1%), with a stronger effect observed at the glans (Figure 2). MC was associated with a significantly decreased risk of prevalent HPV infections at the glans (RR 0·57, 95% CI 0·39–0·82, I^2^=82·2%), but not at the shaft or combined sites (RR 0·96, 95% CI 0·92–1·01, I^2^=0·0%). Findings were similar when stratifying by hrHPV and lrHPV types (Supplementary Figures 1 and 2).

**Figure 2:**
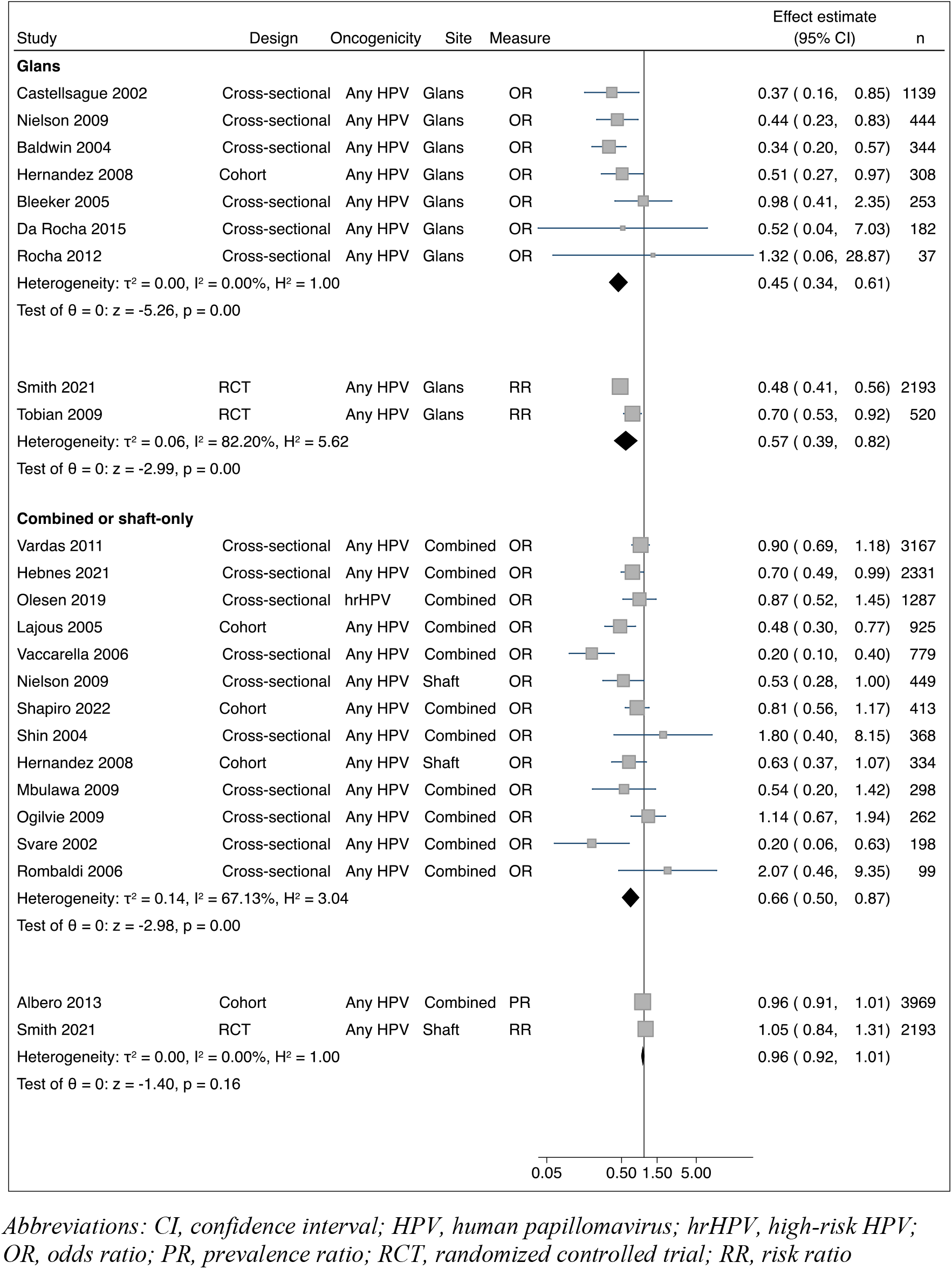
Studies of male circumcision and HPV prevalence in males by sampling site.

Nine studies examined the association between MC and HPV incidence in males (Supplementary Table 4), with sample sizes ranging from 210 to 4,033. A significant protective effect of MC was observed for the incidence rate (IRR 0·69, 95% CI 0·57–0·83, I^2^=0·0%) at the glans, but not for the hazard rate at the shaft or combined sites (HR 1·04, 95% CI 0·94–1·16, I^2^=0·0%) (Figure 3). Results were similar when stratifying by HPV oncogenicity (Supplementary Figures 3 and 4).

**Figure 3:**
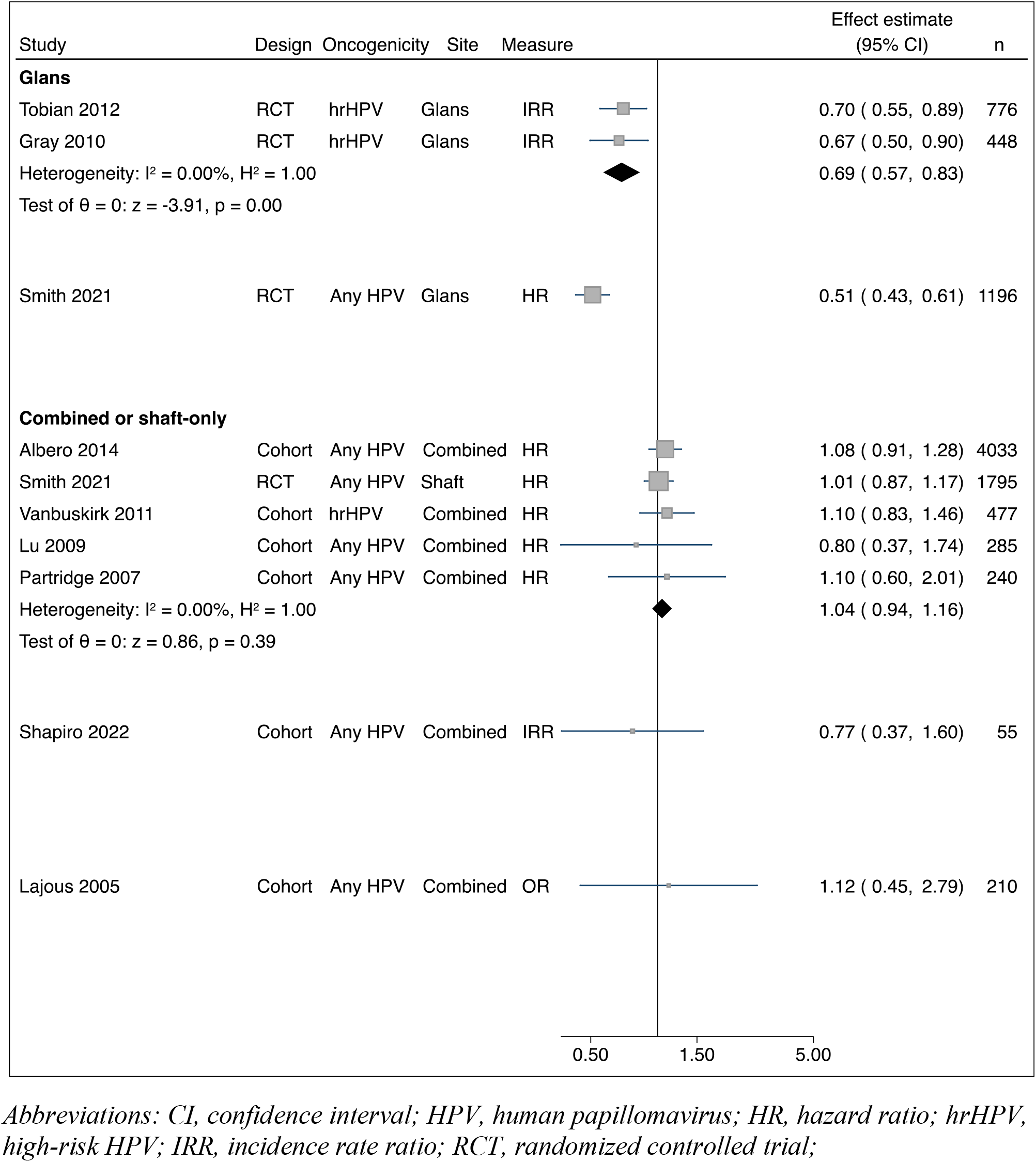
Studies of male circumcision and HPV incidence in males by sampling site.

**Figure 4:**
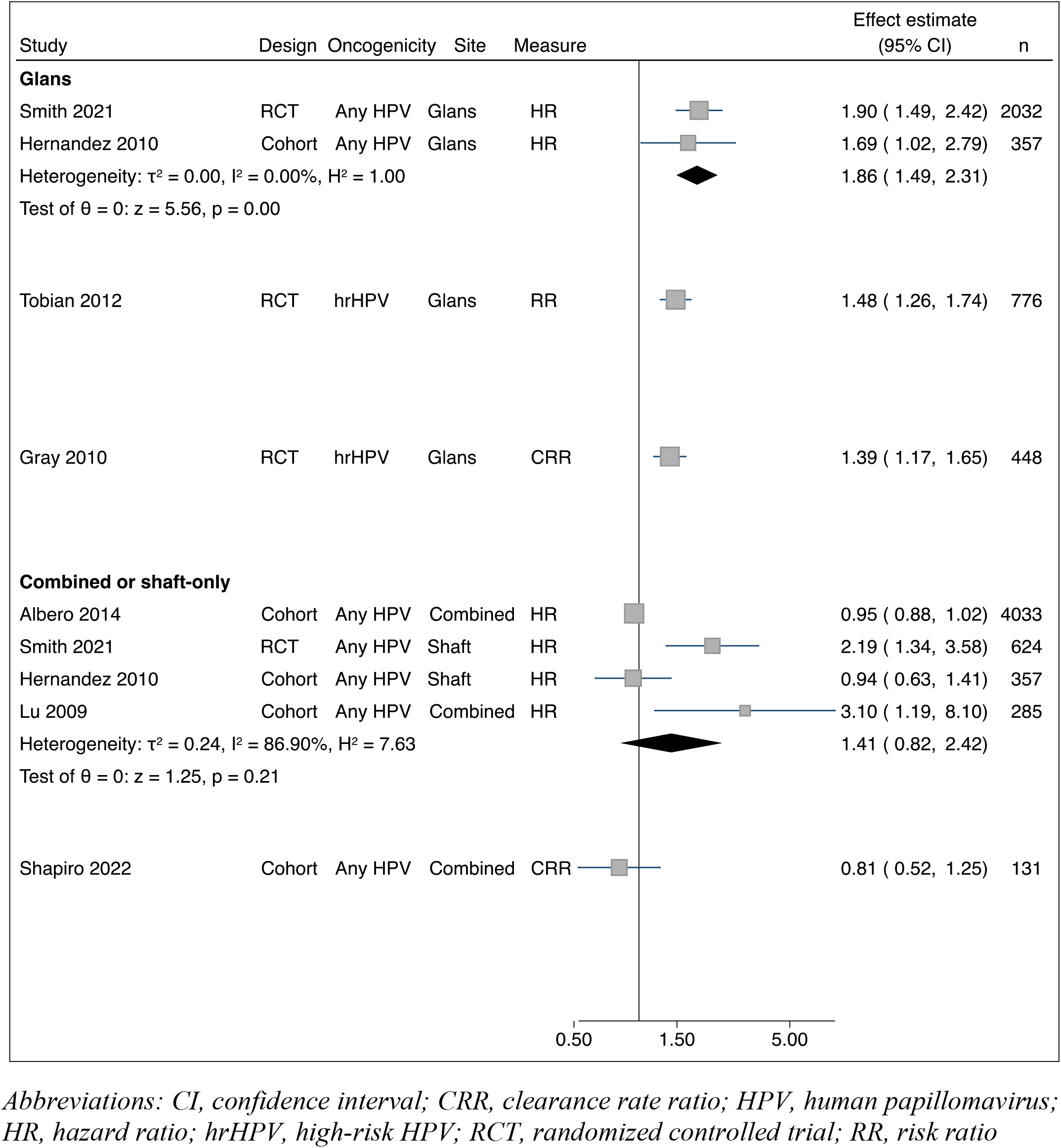
Studies of male circumcision and HPV clearance in males by sampling site.

Seven publications, with sample sizes of 285 to 4,033, examined the association between MC and HPV clearance in males (Supplementary Table 5). Both the risk and hazard rate of HPV infection clearance were significantly increased at the glans of circumcised males (HR 1·86, 95% CI 1·49– 2·31, I^2^=0·0%, RR 1·44, 95% CI 1·28–1·61, I^2^=0·0%) (Figure 5), while the hazard rate was increased at the shaft and combined sites, albeit non-significantly (HR 1·41, 95% CI 0·81–2·42, I^2^=86·9%). Results remained similar when separately examining hrHPV and lrHPV.

**Figure 5:**
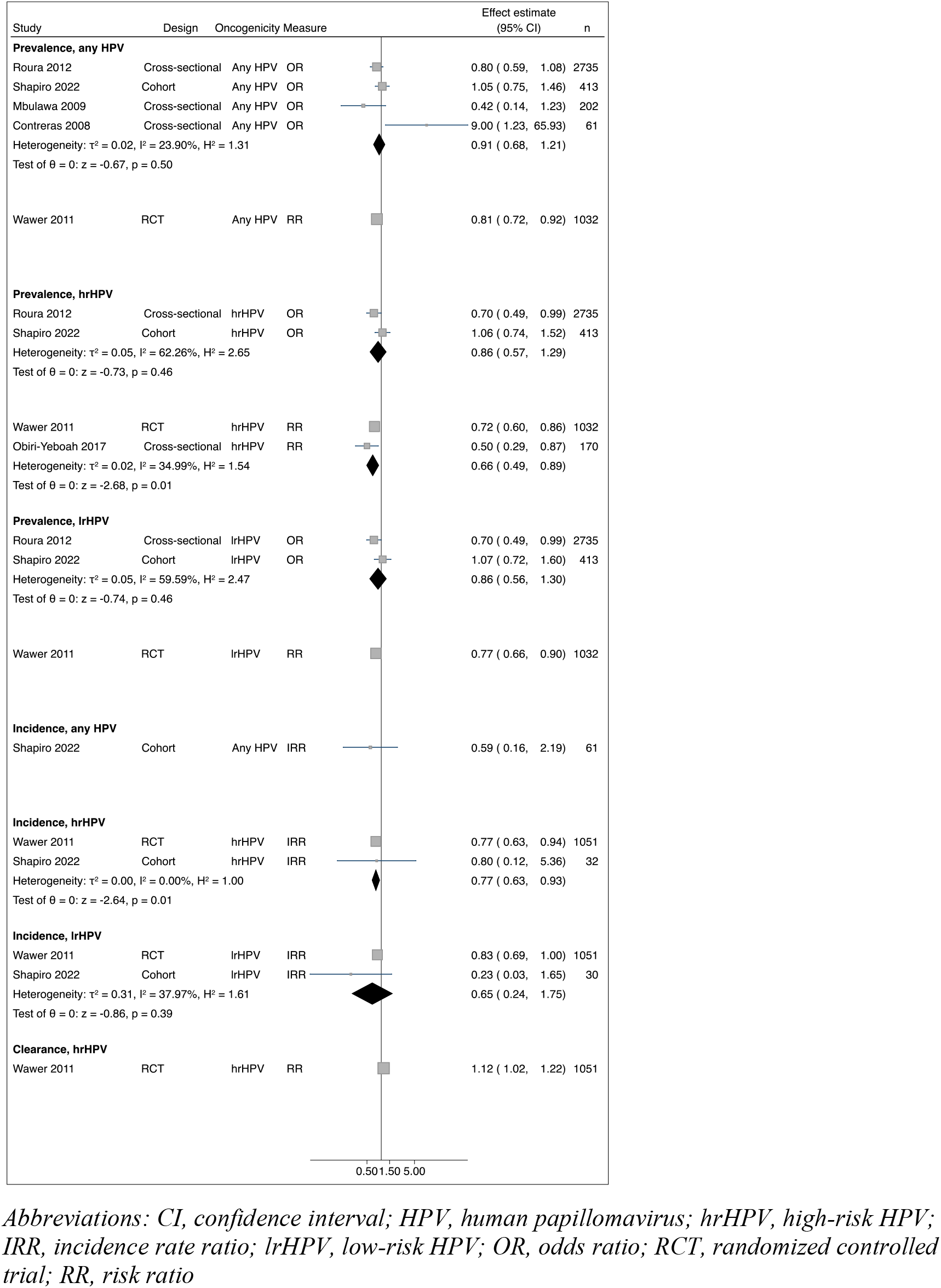
Studies of male circumcision and various HPV outcomes in females.

Six studies examined the association between MC and various HPV outcomes in females (Supplementary Table 6). Sample sizes ranged from 61 to 2,735. All studies assessed the prevalence of HPV infections, while two additionally assessed the acquisition of HPV infections and one assessed the clearance of HPV infections. The risk of prevalent hrHPV infections and the incident rate of hrHPV infections were significantly reduced in female partners of circumcised males (RR 0·66, 95% CI 0·49–0·89, I^2^=35·0%, IRR 0·77, 95% CI 0·63–0·93, I^2^=0·0%) (Figure 5). For all other outcomes, point estimates were protective, but did not reach statistical significance.

We found evidence of an effect modification of the association between MC and HPV prevalence in males by sampling site, with a 32% increase in the protective effect of MC at the glans than at the shaft or combined sites (OR 0·68, 95% CI 0·48–0·98) (Table 2). No effect modification was observed for year of publication, primary study country’s economic development, or whether the study accounted for confounding.

**Table 2.** Meta-regression of studies assessing the association between male circumcision and HPV prevalence in males.

| Potential effect modifier | Number of studies (%) | Univariate |  |
| --- | --- | --- | --- |
|  |  | OR (95% CI) | p-value for modifier |
| Year |  |  |  |
| 2009 or earlier | 15 (62.5) | 1.00 (reference) |  |
| 2010 or later | 9 (37.5) | 1.28 (0.88–1.86) | 0.19 |
| Site, n (%) |  |  |  |
| Combined/shaft-only | 15 (62.5) | 1.00 (reference) |  |
| Glans | 9 (37.5) | 0.68 (0.48–0.98) | 0.04 |
| Economic development, n (%) |  |  |  |
| High-income country | 12 (50.0) | 1.00 (reference) |  |
| Low- or middle-income country | 12 (50.0) | 0.87 (0.61–1.24) | 0.45 |
| Control for confounding, n (%) |  |  |  |
| Yes | 17 (70.8) | 1.00 (reference) |  |
| No | 7 (29.2) | 1.54 (0.89–2.67) | 0.12 |

*Abbreviations: CI, confidence interval; HPV, human papillomavirus; OR, odds ratio*

Most studies were judged to have a low risk of bias (Supplementary Tables 7–9). Restricting to studies with a low risk of bias did not change any findings for studies of prevalence (Supplementary Figure 7), incidence (Supplementary Figure 8), and clearance (Supplementary Figure 9) of HPV infections in males. When stratifying studies of prevalence in males by whether or not they controlled for confounding, we observed that studies that did control for confounding found significantly protective effects of MC at the glans on both the odds and risk scales (Supplementary Figure 10). Studies that did not control for confounding only evaluated the effects of MC at the glans on the odds scale and did not find a protective effect. Studies that controlled for confounding also found significantly protective effects of MC at the shaft and combined sites on the odds scale, but not the risk scale, and studies that did not control for confounding did not find any protective effect of MC at the shaft or combined sites. Excluding any given study of prevalence in males reporting an OR did not significantly change the pooled estimate (Supplementary Figure 11), and we did not observe evidence of publication bias for these studies (Supplementary Figure 12, p value for Egger test 0·95). Publication bias could not be assessed for other outcomes due to the limited number of studies that used the same effect measure.

## DISCUSSION

Findings of this meta-analysis suggest that MC results in reduced prevalent and incident HPV infections and increased clearance of HPV infections at the glans penis, as well as reduced prevalent infections at the shaft. Protection may also be conferred to the female sexual partners of circumcised males. Our findings of the protective effect of MC against various HPV infection outcomes are consistent with those of previous reviews.^10-14^ However, our analysis of the varying effect of MC at different anatomical sites of the penis and the use of a meta-regression approach to assess for effect modification have not been done before. To the best of our knowledge, our analysis seems to be the first to include both males and females in the same review.

Infections with hrHPV are of most clinical relevance, as persistent infection with hrHPV is a necessary cause of cervical cancer and is associated with various anogenital cancers.^2-4^ All estimates for the association between MC and hrHPV infection prevalence, incidence, and clearance found that MC had a significantly protective effect at the glans, and either a protective effect or no effect at the shaft. MC was not found to be a risk factor for HPV infections in any of our meta-analyses.

We included several publications that were not part of the most recently published systematic reviews on the topic: in males, we included an additional nine records of prevalence,^8,24,29,31,36,40,42,45,51^ three of incidence,^8,45,48^ and four of clearance^8,32,45,48^ that were not included in Zhu’s 2017 review and meta-analysis^14^. In females, we added three records^28,36,45^ that were absent in Morris’ 2019 review.^11^ The addition of new records did not result in different conclusions than those of previous reviews, but rather provided further and more detailed evidence for the same interpretations, especially for the varying levels of protection MC confers at different anatomical sites of the penis.

The biological mechanism by which MC is suggested to protect against HPV infections is still unclear; the prevailing theories suggesting differences in keratinization and in the local immune environment of the penis as plausible. It was originally thought that the glans of the circumcised penis is more keratinized than that of the uncircumcised penis^53^ and less vulnerable to the acquisition of sexually transmitted infections during sexual intercourse. However, anatomic and histological studies have failed to find consistent results on the differences in keratinization between the glans of circumcised and uncircumcised males.^54^ MC has also been postulated to change the local immune environment of the penis through changes in the microbiome and immune cell density. Removal of the foreskin eliminates the anaerobic environment of the preputial cavity.^55^ The Ugandan trial of MC found that circumcised males had a decreased total bacterial load and reduced biodiversity in their microbiota,^56^ whereas a 2017 study of 51 females showed that those who were HPV-positive were more likely to have a diverse array of facultative and strict anaerobic bacteria in their vaginal microbiome.^57^ MC may protect against HPV by reducing the diversity of anaerobic bacteria in the penile microbiota. Finally, different anatomical sites of the penis have different distributions of immune cells.^58^ The removal of the foreskin and the immune cells within it may result in different cytokine environments and inflammatory responses to pathogen entry, both of which are associated with the risk of HPV infections.^54,59-61^ Our review had many strengths. We searched a diverse array of databases and validated our search strategy with a librarian. We did not apply study design or language restrictions and we included both males and females, multiple HPV infection-related outcomes, different HPV risk groupings, and different anatomical sampling sites.

Our review also had several limitations. We included the term “circumcision” in our search strategy and may not have captured records that measured MC and HPV infection without directly assessing their association. We were unable to consider other factors that may play a role in MC’s association with HPV infection, such as method of MC, whether MC was performed before or after sexual debut, and number of sexual partners, as these variables were not collected in the vast majority of the included studies. Only three of the 25 unique study populations included in our review came from RCTs, which limited our ability to assess causality. However, it is noteworthy that all RCTs assessing HPV infections in males^8,9,30,48^ found a protective effect of MC at the glans for prevalence, incidence and clearance of all HPV types, including hrHPV, and all estimates but one were statistically significant.

In conclusion, results from our systematic review and meta-analysis support that MC protects against HPV infections in a diverse population of males, particularly at the glans, and that protection may be passed on to female partners. MC may be a viable preventive strategy for HPV infections, especially in regions with a high burden of HPV-associated cancers and where the HPV vaccine is not commercially available.

## Data Availability

All data used in the systematic review, meta-analysis, and meta-regression are available in the supplementary material.

## Contributors

SBS and ELF conceptualized the project. SBS and CL conducted the literature search, screening, and data extraction. CL and SBS performed data analysis. SBS drafted the manuscript. CL, MZ, and ELF critically reviewed the manuscript. ELF and MZ provided supervision and guidance. All authors had access to all the data in the study and had final responsibility for the decision to submit for publication. Both SBS and CL directly accessed and verified the underlying data reported in the manuscript.

## Data sharing

All study-level data used in this study are available in the Supplementary Material.

## Acknowledgments

We would like to thank the librarian Genevieve Gore at McGill University for her assistance in developing and validating our search strategy.

