## Supplementary Material for "Association between male circumcision and human papillomavirus infection in males and females: a systematic review, meta-analysis, and meta-regression"

#### Supplementary Table 1: Search strategy by database

| MEDLINE | Embase | Scopus | Cochrane | LILACS | ProQuest |
|---|---|---|---|---|---|
| 1. Circumcision, Male/<br>2. (circumcis* OR uncircumcis*).ti,ab,kf.<br>3. 1 OR 2<br>4. Papillomavirus Infections/<br>5. papillomaviridae/ OR exp alphapapillomavirus/<br>6. (hpv OR papillomavir* OR papilloma vir*).ti,ab,kf.<br>7. 4 OR 5 OR 6<br>8. 3 AND 7 | 1. circumcision/<br>2. circumcis*.ti,ab,kw<br>3. uncircumcis*.ti,ab,kw<br>4. 1 OR 2 OR 3<br>5. papillomavirus infection/<br>6. papillomaviridae/<br>7. Papilloma virus/<br>8. exp Alphapapillomavirus/<br>9. hpv*.ti,ab,kw<br>10. papillomavir*.ti,ab,kw<br>11. "papilloma vir*".ti,ab,kw<br>12. alphapapilloma*.ti,ab,kw<br>13. "alpha papilloma*".ti,ab,kw<br>14. 5 OR 6 OR 7 OR 8 OR 9 OR 10 OR 11 OR 12 OR 13<br>15. 4 AND 14 | (TITLE-ABS-KEY(circumcis*) OR TITLE-ABS-KEY(uncircumcis*))<br><br>AND<br><br>(TITLE-ABS-KEY(hpv*) OR TITLE-ABS-KEY(papillomavir*) OR TITLE-ABS-KEY(papilloma vir*) OR TITLE-ABS-KEY(alphapapilloma*) OR TITLE-ABS-KEY(alpha papilloma*)) | 1. [mh ^"Circumcision, male"]<br>2. circumcis*:ti,ab,kw<br>3. uncircumcis*:ti,ab,kw<br>4. #1 OR #2 OR #3<br>5. [mh ^"Papillomavirus Infections"]<br>6. [mh ^"Papillomaviridae"]<br>7. [mh "alphapapillomavirus"]<br>8. hpv*:ti,ab,kw<br>9. papillomavir*:ti,ab,kw<br>10. papilloma vir*:ti,ab,kw<br>11. alphapapilloma*:ti,ab,kw<br>12. alpha papilloma*:ti,ab,kw<br>13. #5 OR #6 OR #7 OR #8 OR #9 OR #10 OR #11 OR #12<br>14. #4 AND #13 | ["papillomavirus infections" or "papillomaviridae" or "alphapapillomavirus" (in DeCS category)] OR[hpv$ or papillomavir$ or "papilloma vir$" or alphapapilloma$ or "alpha papilloma$" (in words)]<br><br>AND<br><br>[circumcis$ or uncircumcis$ (in words)] | (circumcis* OR uncircumcis*)<br><br>AND<br><br>(HPV OR human papillomavir* OR papillomavir* OR papilloma vir* OR papillomaviridae) |

#### Supplementary Table 2: Reason for excluding studies at full-text screening

| Reason | Study |
|---|---|
| Reporting on already included cohort | Auvert B, Lissouba P, Cutler E, Zarca K, Puren A, Taljaard D. Association of oncogenic and nononcogenic human papillomavirus with HIV incidence. Journal of Acquired Immune Deficiency Syndromes: JAIDS 2010; 53(1): 111–6. |
| | Backes DM, Bleeker MCG, Meijer CJLM, et al. Male circumcision is associated with a lower prevalence of human papillomavirus-associated penile lesions among Kenyan men. Int J Cancer 2012; 130(8): 1888–97. |
| | Backes DM, Smith JS. Natural history of human papillomavirus infection among young men from Kisumu, Kenya. 2010; (3418481): 106. |
| | Baldwin SB, Wallace DR, Papenfuss MR, et al. Human papillomavirus infection in men attending a sexually transmitted disease clinic. J Infect Dis 2003; 187(7): 1064–70. |
| | Daugherty M, Byler T. HPV prevalence in males in the United States from penile swabs: Results from NHANES. J Urol 2017; 197: e137-e8. |
| | Davis M, Grabowski M, Gravitt P, et al. Male circumcision reduces high-risk human papillomavirus (HPV) viral shedding in female partners in Rakai, Uganda. Gynecol Oncol 2013; 130: e26-e7. |
| | Edelstein ZR, Carter JJ, Garg R, et al. Serum antibody response following genital alpha9 human papillomavirus infection in young men. J Infect Dis 2011; 204(2): 209–16. |
| | Giuliano AR, Lazcano E, Villa LL, et al. Circumcision and sexual behavior: factors independently associated with human papillomavirus detection among men in the HIM study. Int J Cancer 2009; 124(6): 1251–7. |
| | Grabowski MK, Gravitt PE, Gray RH, et al. Trends and Determinants of Human Papillomavirus Concordance Among Human Immunodeficiency Virus-Positive and -Negative Heterosexual Couples in Rakai, Uganda. J Infect Dis 2017; 215(5): 772–80. |
| | Grabowski MK, Kong X, Gray RH, et al. Partner Human Papillomavirus Viral Load and Incident Human Papillomavirus Detection in Heterosexual Couples. J Infect Dis 2016; 213(6): 948–56. |
| | Nielson CM, Flores R, Harris RB, et al. Human papillomavirus prevalence and type distribution in male anogenital sites and semen. Cancer Epidemiol Biomarkers Prev 2007; 16(6): 1107–14. |
| | Olesen TB, Mwaiselage J, Iftner T, et al. Risk factors for genital human papillomavirus among men in Tanzania. J Med Virol 2017; 89(2): 345–51. |
| | Onywera H, Williamson AL, Cozzuto L, et al. The penile microbiota of Black South African men: relationship with human papillomavirus and HIV infection. BMC Microbiol 2020; 20(1): 78. |
| | Senkomago V, Backes DM, Hudgens MG, et al. Acquisition and persistence of human papillomavirus 16 (HPV-16) and HPV-18 among men with high-HPV viral load infections in a circumcision trial in Kisumu, Kenya. J Infect Dis 2015; 211(5): 811–20. |
| | Senkomago V, Smith JS. The role of human papillomavirus (HPV) viral load in penile HPV infection and clearance among young Kenyan men. 2013; (3606765): 91. |
| | Smith JS, Moses S, Hudgens MG, et al. Increased risk of HIV acquisition among Kenyan men with human papillomavirus infection. J Infect Dis 2010; 201(11): 1677–85. |
| | Tobian AAR, Grabowski MK, Kigozi G, et al. High-risk human papillomavirus prevalence is associated with HIV infection among heterosexual men in Rakai, Uganda. Sexually Transmitted Infections 2013; 89(2): 122–7. |
| | Tobian AAR, Kong X, Gravitt PE, et al. Male circumcision and anatomic sites of penile high-risk human papillomavirus in Rakai, Uganda. Int J Cancer 2011; 129(12): 2970–5. |
| Participants have HPV-associated lesions | Aung MT, Soe MY, Mya WW. Study on risk factors for cervical carcinoma at Central Womens Hospital, Yangon, Myanmar. BJOG 2012; 119: 124. |
| | Aynaud O. Intra-epithelial neoplasia of the penis: Virology, histology and clinical aspects. [French]. Andrologie 1994; 4(4): 440–4. |
| | Aynaud O, Huynh B, Bergeron C. Prevalence of HPV-induced lesions in men: A study from 246 heterosexual couples. [French]. Gynecologie Obstetrique et Fertilite 2012; 40(7): 406–10. |
| | Aynaud O, Ionesco M, Barrasso R. Penile intraepithelial neoplasia. Specific clinical features correlate with histologic and virologic findings. Cancer 1994; 74(6): 1762–7. |
| | Aynaud O, Piron D, Bijaoui G, Casanova JM. Developmental factors of urethral human papillomavirus lesions: correlation with circumcision. BJU Int 1999; 84(1): 57–60. |
| | Aynaud O, Poveda JD, Huynh B, Guillemotonia A, Barrasso R. Frequency of herpes simplex virus, cytomegalovirus and human papillomavirus DNA in semen. Int J STD AIDS 2002; 13(8): 547–50. |
| | Chalya PL, Rambau PF, Masalu N, Simbila S. Ten-year surgical experiences with penile cancer at a tertiary care hospital in northwestern Tanzania: a retrospective study of 236 patients. World J Surg Oncol 2015; 13: 71. |
| | Chaux A, Netto GJ, Rodriguez IM, et al. Epidemiologic profile, sexual history, pathologic features, and human papillomavirus status of 103 patients with penile carcinoma. World J Urol 2013; 31(4): 861–7. |
| | D'Hauwers KWM, Depuydt CE, Bogers JJ, et al. Human papillomavirus, lichen sclerosus and penile cancer: A study in Belgium. Vaccine 2012; 30(46): 6573–7. |
| | de Andrade RT, Arcoverde MAL, Vilar FO, Santos Jr MW, Filho NTP, Lima SVC. Clinical epidemiologic study of penile cancer in the state of Pernambuco, Brazil. UroToday International Journal 2012; 5(1). |
| | De Sousa IDB, Vidal FCB, Branco Vidal JPC, De Mello GCF, Do Desterro Soares Brandao Nascimento M, Brito LMO. Prevalence of human papillomavirus in penile malignant tumors: Viral genotyping and clinical aspects. BMC Urol 2015; 15(1): 13. |
| | do Carmo Alves Martins V, Cunha IW, Figliuolo G, et al. Presence of HPV with overexpression of p16INK4a protein and EBV infection in penile cancer-A series of cases from Brazil Amazon. PLoS ONE [Electronic Resource] 2020; 15(5): e0232474. |

| | |
|---|---|
| | Franklin A, Koehne E, Haden T, et al. A review of the stage recurrence treatment and outcomes in penile cancer at a tertiary referral center. Journal of Clinical Oncology Conference 2016; 34(2). |
| | Katelaris PM, Cossart YE, Rose BR, et al. Human papillomavirus: the untreated male reservoir. J Urol 1988; 140(2): 300–5. |
| | Kravvas G, Ge L, Ng J, et al. The management of penile intraepithelial neoplasia (PeIN): clinical and histological features and treatment of 345 patients and a review of the literature. Journal of Dermatological Treatment 2020: 1–16. |
| | Maden C, Sherman KJ, Beckmann AM, et al. History of circumcision, medical conditions, and sexual activity and risk of penile cancer. J Natl Cancer Inst 1993; 85(1): 19–24. |
| | Madsen BS, van den Brule AJ, Jensen HL, Wohlfahrt J, Frisch M. Risk factors for squamous cell carcinoma of the penis--population-based case-control study in Denmark. Cancer Epidemiol Biomarkers Prev 2008; 17(10): 2683–91. |
| | Malek RS, Goellner JR, Smith TF, Espy MJ, Cupp MR. Human papillomavirus infection and intraepithelial, in situ, and invasive carcinoma of penis. Urology 1993; 42(2): 159–70. |
| | Mallon E, Hawkins D, Dinneen M, et al. Circumcision and genital dermatoses. Arch Dermatol 2000; 136(3): 350–4. |
| | Mentrikoski MJ, Stelow EB, Culp S, Frierson Jr HF, Cathro HP. Histologic and immunohistochemical assessment of penile carcinomas in a North American population. The American journal of surgical pathology 2014; 38(10): 1340–8. |
| | Muller EE, Chirwa TF, Lewis DA. Human papillomavirus (HPV) infection in heterosexual South African men attending sexual health services: associations between HPV and HIV serostatus. Sexually Transmitted Infections 2010; 86(3): 175–80. |
| | Park SJ, Seo J, Ha SH, Jung GW. Prevalence and determinants of high-risk human papillomavirus infection in male genital warts. Korean Journal of Urology 2014; 55(3): 207–12. |
| | Porter WM, Francis N, Hawkins D, Dinneen M, Bunker CB. Penile intraepithelial neoplasia: clinical spectrum and treatment of 35 cases. Br J Dermatol 2002; 147(6): 1159–65. |
| | Santos-Cortes J, Shapiro EY, Ghavamian R. Penile carcinoma: A single institution experience. Central European Journal of Urology 2010; 63(3): 117–20. |
| | Seo J, Park S, Jung G. The prevalence and risk factors of high-risk human papillomavirus types in genital condylomata acuminata of Korean male. Urology 2012; 80(3): S155. |
| No genital sample taken | Aung ET, Fairley CK, Tabrizi SN, et al. Detection of human papillomavirus in urine among heterosexual men in relation to location of genital warts and circumcision status. Sexually Transmitted Infections 2018; 94(3): 222–5. |
| | Daling JR, Madeleine MM, Johnson LG, et al. Penile cancer: importance of circumcision, human papillomavirus and smoking in in situ and invasive disease. Int J Cancer 2005; 116(4): 606–16. |
| | Dickson NP, Ryding J, van Roode T, et al. Male circumcision and serologically determined human papillomavirus infection in a birth cohort. Cancer Epidemiol Biomarkers Prev 2009; 18(1): 177–83. |
| | Edmonds EVJ, Hunt S, Hawkins D, Dinneen M, Francis N, Bunker CB. Clinical parameters in male genital lichen sclerosus: A case series of 329 patients. J Eur Acad Dermatol Venereol 2012; 26(6): 730–7. |
| | Homfray V, Tanton C, Miller RF, et al. Male Circumcision and STI Acquisition in Britain: Evidence from a National Probability Sample Survey. PLoS ONE [Electronic Resource] 2015; 10(6): e0130396. |
| | Kolawole OM, Olatunji KT, Durowade KA, Adeniyi AA, Omokanye LO. Prevalence, risk factors of human papillomavirus infection and papanicolaou smear pattern among women attending a tertiary health facility in south-west Nigeria. TAF Preventive Medicine Bulletin 2015; 14(6): 451–7. |
| | Varda BK, Cheng PJ, Cendron M, Chang SL. Circumcision status and HIV, HSV, and HPV: A contemporary analysis of U.S. men using the national health and nutrition survey (NHANES). J Urol 2014; 191(4): e161. |
| No exposure or outcome data | Afonso LA, Cordeiro TI, Carestiato FN, Ornellas AA, Alves G, Cavalcanti SMB. High Risk Human Papillomavirus Infection of the Foreskin in Asymptomatic Men and Patients with Phimosis. J Urol 2016; 195(6): 1784–9. |
| | Garolla A, Pizzol D, Vasoin F, Barzon L, Bertoldo A, Foresta C. Counseling Reduces HPV Persistence in Coinfected Couples. J Sex Med 2014; 11(1): 127–35. |
| | Kasap B, Yetimalar H, Keklik A, Yildiz A, Cukurova K, Soylu F. Prevalence and risk factors for human papillomavirus DNA in cervical cytology. Eur J Obstet Gynecol Reprod Biol 2011; 159(1): 168–71. |
| | Kim J, Kim BK, Lee CH, Seo SS, Park SY, Roh JW. Human papillomavirus genotypes and cofactors causing cervical intraepithelial neoplasia and cervical cancer in Korean women. Int J Gynecol Cancer 2012; 22(9): 1570–6. |
| | Kjaer SK, de Villiers EM, Dahl C, et al. Case-control study of risk factors for cervical neoplasia in Denmark. I: Role of the "male factor" in women with one lifetime sexual partner. Int J Cancer 1991; 48(1): 39–44. |
| | Lorenzon L, Terrenato I, Dona MG, et al. Prevalence of HPV infection among clinically healthy Italian males and genotype concordance between stable sexual partners. J Clin Virol 2014; 60(3): 264–9. |
| | Viladoms Fuster JM, Leira Juanos J. [Human papilloma virus in the male]. Actas Urol Esp 1989; 13(5): 343–6. |
| Outcome not of interest | Burchell AN, Coutlee F, Tellier P, Hanley J, Franco EL. Transmission of human papillomavirus infections. Sexually Transmitted Infections 2011; 87: A14. |
| | Davis MA, Gray RH, Grabowski MK, et al. Male circumcision decreases high-risk human papillomavirus viral load in female partners: A randomized trial in Rakai, Uganda. Int J Cancer 2013; 133(5): 1247–52. |
| | Grabowski MK, Gray RH, Serwadda D, et al. High-risk human papillomavirus viral load and persistence among heterosexual HIV-negative and HIV-positive men. Sexually Transmitted Infections 2014; 90(4): 337–43. |
| | Lazcano-Ponce E, Sudenga SL, Nelson Torres B, et al. Incidence of external genital lesions related to human papillomavirus among Mexican men. A cohort study. Salud Publica Mex 2018; 60(6): 633–44. |
| | Rositch AF, Mao L, Hudgens MG, et al. Risk of HIV acquisition among circumcised and uncircumcised young men with penile human papillomavirus infection. AIDS 2014; 28(5): 745–52. |
| | Schabath MB, Villa LL, Lazcano-Ponce E, Salmeron J, Quiterio M, Giuliano AR. Smoking and human papillomavirus (HPV) infection in the HPV in men (HIM) study. Cancer Epidemiology Biomarkers and Prevention 2012; 21(1): 102–10. |

| | |
|---|---|
| | Sichero L, Pierce Campbell CM, Fulp W, et al. High genital prevalence of cutaneous human papillomavirus DNA on male genital skin: The HPV infection in men study. BMC Infect Dis 2014; 14(1). |
| No associated full-text article | Raghavendran A, George AJ, Manojkumar R, Devasia A, Abraham P. HPV in asymptomatic men in the Indian sub-continent. BMC Infectious Diseases Conference: International Science Symposium on HIV and Infectious Diseases, ISSHID 2019; 20. |
| | Wei FX, Guo M, Ma XJ, et al. [The impact of male circumcision on the natural history of genital HPV infection: a prospective cohort study]. Chung-Hua Yu Fang i Hsueh Tsa Chih [Chinese Journal of Preventive Medicine] 2018; 52(5): 486–92. |
| Could not obtain data | Auvert B, Sobngwi-Tambekou J, Cutler E, et al. Effect of male circumcision on the prevalence of high-risk human papillomavirus in young men: results of a randomized controlled trial conducted in Orange Farm, South Africa. J Infect Dis 2009; 199(1): 14–9. |
| | Berenson AB, Hirth JM, Chang M. Prevalence of genital human papillomavirus by age and race/ ethnicity among males. Clin Infect Dis 2021; 13: 13. |
| | Brinton LA, Reeves WC, Brenes MM, et al. The male factor in the etiology of cervical cancer among sexually monogamous women. Int J Cancer 1989; 44(2): 199–203. |
| | Huang LL, Deng JH, Shi H, et al. [Circumcision reduces the incidence of human papillomavirus infection in men]. Zhong Hua Nan Ke Xue 2018; 24(4): 327–30. |
| | Ng'ayo MO, Bukusi E, Rowhani-Rahbar A, et al. Epidemiology of human papillomavirus infection among fishermen along Lake Victoria Shore in the Kisumu District, Kenya. Sex Transm Infect 2008; 84(1): 62–6. |
| | Tarnaud C, Lissouba P, Cutler E, Puren A, Taljaard D, Auvert B. Association of low-risk human papillomavirus infection with male circumcision in young men: results from a longitudinal study conducted in Orange Farm (South Africa). Infect Dis Obstet Gynecol 2011; 2011: 567408. |
| | Tobian AA, Kigozi G, Gravitt PE, et al. Human papillomavirus incidence and clearance among HIV-positive and HIV-negative men in sub-Saharan Africa. AIDS 2012; 26(12): 1555–65. |
| | Weaver BA, Feng Q, Holmes KK, et al. Evaluation of genital sites and sampling techniques for detection of human papillomavirus DNA in men. J Infect Dis 2004; 189(4): 677–85. |

#### Supplementary Table 3: Studies of MC and HPV prevalence in males by HPV risk grouping

| First author & year | Study design | HPV types | Age range at baseline (years) | Circumcision prevalence (%) | HPV prevalence at baseline (%) | Sampling site | Sample size | Effect estimate: OR (95% CI) | Covariate adjustment |
|---|---|---|---|---|---|---|---|---|---|
| *Any HPV* | | | | | | | | | |
| Albero 2013 | Cohort[b] | 37 types: 16, 18, 31, 33, 35, 39, 45, 51, 52, 56, 58, 59, 68, 6, 11, 26, 40, 42, 53, 54, 55, 61, 62, 64, 66, 67, 69, 70, 71, 72, 73, 81, 82, 82/IS39, 83, 84, 89 | 18–70 | 35·9 | 66·7 | Combined | 3969 | PR 0·96 (0·91–1·01) | Race, marital status, lifetime female sexual partners, female sexual partners in past 3–6 months, male anal sexual partners in the past 3 months |
| Vardas 2011 | Cross-sectional | 14 types: 6, 11, 16, 18, 31, 33, 35, 39, 45, 51, 52, 56, 58, 59 | 15–24 | 36·4 | 21·2 | Combined | 3167 | 0·9 (0·7–1·2) | Geographic area of residence, age, tobacco use, condom use, age at first sexual intercourse with a male partner, number of lifetime sexual partners, number of new partners in the past 6 months |
| Hebnes 2021 | Cross-sectional | 24 types: 16, 18, 31, 33, 35, 39, 45, 51, 52, 53, 56, 58, 59, 66, 68, 6, 11, 40, 42, 43, 44, 54, 70, 74 | 18–59 | 5·4 | 41·7 | Combined | 2331 | 0·7 (0·5–1·0) | Age, lifetime number of female sex partners and age at first sexual intercourse with a woman as continuous variables, time since last sexual intercourse as a categorical variable |
| Smith 2021 | RCT[c] | 44 types: 6, 11, 16, 18, 26, 30, 31, 32, 33, 34, 35, 39, 40, 42, 43, 44, 45, 51, 52, 53, 54, 55, 56, 57, 58, 59, 61, 64, 66, 67, 68, 69, 70, 71, 72, 73, 81, 82/MM4, 82/IS39, 83, 84, 85, 86, 89, JC9710 | 18–24 | 50·0 | 50[d] | Combined | 2193 | PR 0·57 (0·49–0·67) | None |
| Smith 2021 | RCT[c] | 44 types: 6, 11, 16, 18, 26, 30, 31, 32, 33, 34, 35, 39, 40, 42, 43, 44, 45, 51, 52, 53, 54, 55, 56, 57, 58, 59, 61, 64, 66, 67, 68, 69, 70, 71, 72, 73, 81, 82/MM4, 82/IS39, 83, 84, 85, 86, 89, JC9710 | 18–24 | 50·0 | 50[d] | Glans | 2193 | PR 0·48 (0·41–0·56) | None |
| Smith 2021 | RCT[c] | 44 types: 6, 11, 16, 18, 26, 30, 31, 32, 33, 34, 35, 39, 40, 42, 43, 44, 45, 51, 52, 53, 54, 55, 56, 57, 58, 59, 61, 64, 66, 67, 68, 69, 70, 71, 72, 73, 81, 82/MM4, 82/IS39, 83, 84, 85, 86, 89, JC9710 | 18–24 | 50·0 | 50[d] | Shaft | 2193 | PR 1·05 (0·84–1·31) | None |
| Castellsague 2002 | Cross-sectional | 6 types: 6, 11, 16, 18, 31, 33 | NR | 25·6 | 16 | Glans | 1139 | 0·37 (0·16–0·85) | Age, study location, level of education, age at first sexual intercourse, lifetime number of |

| | | | | | | | | | |
|---|---|---|---|---|---|---|---|---|---|
| | | | | | | | | | sexual partners, frequency of genital washing after sex |
| Lajous 2005 | Cohort[b] | 27 types: 16, 18, 31, 33, 35, 39, 45, 51, 52, 53, 56, 58, 59, 68, 73, 82, 6, 11, 26, 40, 42, 54, 55, 57, 66, 83, 84 | 16–40 | 10·3 | 44·6 | Combined | 925 | 0·48 (0·30–0·77) | Age, SES, lifetime number of partners |
| Vaccarella 2006 | Cross-sectional | 35 types: 16, 18, 31, 33, 35, 39, 45, 51, 52, 56, 58, 59, 68, 73, 82, 6, 11, 26, 40, 42, 53, 54, 55, 61, 62, 64, 66, 67, 69, 70, 71, 72, 81, 83, 84 | <25–≥45 | 31·7 | 8·7 | Combined | 779 | 0·2 (0·1–0·4) | Age group, lifetime number of sexual partners |
| Tobian 2009 | RCT[c] | 37 types: 16, 18, 31, 33, 35, 39, 45, 51, 52, 56, 58, 59, 66, 68, 6, 11, 26, 40, 42, 53, 54, 55, 61, 62, 64, 67, 69, 70, 71, 72, 73, 81, 82, 82/IS39, 83, 84, 89 | 15–49 | 44·8 | 62·2 | Glans | 520 | RR 0·70 (0·53–0·91) | None |
| Nielson 2009 | Cross-sectional | 37 types: 16, 18, 31, 33, 35, 39, 45, 51, 52, 56, 58, 59, 68, 6, 11, 26, 40, 42, 53, 54, 55, 61, 62, 64, 66, 67, 69, 70, 71, 72, 73, 81, 82, 82/IS39, 83, 84, 89 | 18–40 | 84·1 | 47·5 | Combined | 421 | 0·68 (0·36–1·27) | Date of analysis, smoking status, lifetime number of female sex partners, condom use in the past 3 months |
| Nielson 2009 | Cross-sectional | 37 types: 16, 18, 31, 33, 35, 39, 45, 51, 52, 56, 58, 59, 68, 6, 11, 26, 40, 42, 53, 54, 55, 61, 62, 64, 66, 67, 69, 70, 71, 72, 73, 81, 82, 82/IS39, 83, 84, 89 | 18–40 | 84·1 | 47·5 | Glans | 444 | 0·44 (0·23–0·82) | Date of analysis, smoking status, lifetime number of female sex partners, condom use in the past 3 months |
| Nielson 2009 | Cross-sectional | 37 types: 16, 18, 31, 33, 35, 39, 45, 51, 52, 56, 58, 59, 68, 6, 11, 26, 40, 42, 53, 54, 55, 61, 62, 64, 66, 67, 69, 70, 71, 72, 73, 81, 82, 82/IS39, 83, 84, 89 | 18–40 | 84·1 | 47·5 | Shaft | 449 | 0·53 (0·28–0·99) | Date of analysis, smoking status, lifetime number of female sex partners, condom use in the past 3 months |
| Shapiro 2022 | Cohort[b] | 36 types: 6, 11, 16, 18, 26, 31, 33, 34, 35, 39, 40, 42, 44, 45, 51, 52, 53, 54, 56, 58, 59, 61, 62, 66, 67, 68, 69, 70, 71, 72, 73, 81, 82, 83, 84, 89 | 17–45 | 47·2 | 54·6 | Combined | 413 | 0·81 (0·56–1·16) | Propensity score (male's and female's age, region born, race, educational attainment, smoking history, same-sex partner history, number of sexual partners, number of vaginal sexual partners, age at first sex; female's vaccination against HPV; couple's marital status, condom use, frequency of sex, concurrent partner at future visit) |
| Shin 2004 | Cross-sectional | 25 types: 16, 18, 31, 33, 35, 39, 45, 51, 52, 56, 58, 59, 66, 68/73, 6, 11, 34, 40, 42, 43, 44, 53, 54, 70, 74 | 18–28 | 88·3 | 8·7 | Combined | 368 | 1·8 (0·4–8·2) | Age, number of lifetime sexual partners |

| | | | | | | | | | |
|---|---|---|---|---|---|---|---|---|---|
| Baldwin 2004 | Cross-sectional | 27 types: 16, 18, 26, 31, 33, 35, 39, 45, 51, 52, 55, 56, 58, 59, 68, 73, 82, 83, 6, 11, 40, 42, 53, 54, 57, 66, 84 | 18–70 | 67·4 | 28·2 | Glans | 344 | 0·34 (0·20–0·57) | Sexual frequency per month, genital warts, condom use in past 3 months, steady partner |
| Hernandez 2008 | Cross-sectional | 37 types: 16, 18, 26, 31, 33, 35, 39, 45, 51–53, 56, 58, 59, 66, 68, 73, 82, 82/IS39, 6, 11, 40, 42, 54, 61, 70, 72, 81, 89, 55, 62, 64, 67, 69, 71, 83, 84 | NR | 77·8 | NR | Combined | 316 | 0·58 (0·30–1·14) | Age, birthplace, race/ethnicity, education level, lifetime number of female sex partners, history of sex with men, age at initial sex, condom use, history of genital warts, history of cigarette smoking |
| Hernandez 2008 | Cross-sectional | 37 types: 16, 18, 26, 31, 33, 35, 39, 45, 51–53, 56, 58, 59, 66, 68, 73, 82, 82/IS39, 6, 11, 40, 42, 54, 61, 70, 72, 81, 89, 55, 62, 64, 67, 69, 71, 83, 84 | NR | 77·8 | NR | Glans | 308 | 0·51 (0·27–0·97) | Age, birthplace, race/ethnicity, education level, lifetime number of female sex partners, history of sex with men, age at initial sex, condom use, history of genital warts, history of cigarette smoking |
| Hernandez 2008 | Cross-sectional | 37 types: 16, 18, 26, 31, 33, 35, 39, 45, 51–53, 56, 58, 59, 66, 68, 73, 82, 82/IS39, 6, 11, 40, 42, 54, 61, 70, 72, 81, 89, 55, 62, 64, 67, 69, 71, 83, 84 | NR | 77·8 | NR | Shaft | 334 | 0·63 (0·42–1·22) | Age, birthplace, race/ethnicity, education level, lifetime number of female sex partners, history of sex with men, age at initial sex, condom use, history of genital warts, history of cigarette smoking |
| Mbulawa 2009 | Cross-sectional | 37 types: 16, 18, 31, 33, 35, 39, 45, 51, 52, 56, 58, 59, 68, 73, 82, 26, 53, 66, 6, 11, 40, 42, 54, 55, 61, 62, 64, 67, 69, 70, 71, 72, 81, 82/IS39, 83, 84, 89 | 20–64 | 93·3 | 43 | Combined | 298 | 0·54 (0·20–1·39)[d] | None |
| Ogilvie 2009 | Cross-sectional | 13 types: 16, 18, 31, 33, 35, 39, 45, 51, 52, 56, 58, 59, 68 | 16–69 | 50·4 | 69·8 | Combined | 262 | 1·14 (0·67–1·94)[d] | None |
| Bleeker 2005 | Cross-sectional | 37 types: 16, 18, 31, 33, 35, 39, 45, 51, 52, 56, 58, 59, 66, 68, 6, 11, 26, 34, 40, 42, 43, 44, 53, 54, 55, 57, 61, 70, 71, 72, 73, 81, 82/MM4, 82/IS39, 83, 84, 89 | 22·5–73·2 | 9·1 | 48·2 | Glans | 253 | 0·98 (0·41–2·36)[d] | None |
| Svare 2002 | Cross-sectional | NR | 18–≥40 | 12·1 | 45 | Combined | 198 | 0·2 (0·06–0·6) | Age, lifetime sex partners, sex partners in past year, genital warts |
| Da Rocha 2015 | Cross-sectional | 11 types: 16, 18, 31, 33, 35, 45, 56, 58, 6, 11, 53 | 18–65 | 3·3 | 16·5 | Glans | 182 | 0·52 (0·02–3·66)[d] | None |
| Rombaldi 2006 | Cross-sectional | 32 types: 6, 11, 13, 16, 18, 26, 31, 32, 33, 34, 35, 39, 40, 42, 44, 45, 51, 52, 53, 54, 55, 56, 57, 58, 59, 61, 62, 64, 66, 67, 68, 69 | ≤19–59 | 10·1 | 54·5 | Combined | 99 | 2·07 (0·51–10·41)[d] | None |
| Rocha 2012 | Cross-sectional | 7 types: 16, 18, 31, 33, 45, 6, 11 | 18–60 | 1·3 | 52·1 | Glans | 37 | 1·32 (0·06–28·7)[d] | None |

| | | | | | | | | | |
|---|---|---|---|---|---|---|---|---|---|
| *High-risk HPV* | | | | | | | | | |
| Albero 2013 | Cohort[b] | 13 types: 16, 18, 31, 33, 35, 39, 45, 51, 52, 56, 58, 59, 68 | 18–70 | 35·9 | 66·7 | Combined | 3969 | PR 0·95 (0·87–1·03) | Race, marital status, lifetime female sexual partners, female sexual partners in past 3–6 months, male anal sexual partners in the past 3 months |
| Smith 2021 | RCT[c] | 14 types: 16, 18, 31, 33, 35, 39, 45, 51, 52, 56, 58, 59, 66, 68 | 18–24 | 50·0 | 50[d] | Combined | 2193 | PRR 0·55 (0·46–0·67) | None |
| Smith 2021 | RCT[c] | 14 types: 16, 18, 31, 33, 35, 39, 45, 51, 52, 56, 58, 59, 66, 68 | 18–24 | 50·0 | 50[d] | Glans | 2193 | PRR 0·46 (0·38–0·57) | None |
| Smith 2021 | RCT[c] | 14 types: 16, 18, 31, 33, 35, 39, 45, 51, 52, 56, 58, 59, 66, 68 | 18–24 | 50·0 | 50[d] | Shaft | 2193 | PRR 1·20 (0·88–1·62) | None |
| Olesen 2019 | Cross-sectional | 13 types: 16, 18, 31, 33, 35, 39, 45, 51, 52, 56, 58, 59, 68 | 14–90 | 91·5 | 17 | Combined | 1287 | 0·87 (0·53–1·47)[d] | None |
| Tobian 2009 | RCT[c] | 14 types: 16, 18, 31, 33, 35, 39, 45, 51, 52, 56, 58, 59, 66, 68 | 15–49 | 44·8 | 62·2 | Glans | 520 | RR 0·65 (0·46–0·90) | Enrollment characteristics, rates of sexual practices, symptoms of sexually transmitted infections |
| Nielson 2009 | Cross-sectional | 13 types: 16, 18, 31, 33, 35, 39, 45, 51, 52, 56, 58, 59, 68 | 18–40 | 84·1 | 47·5 | Combined | 421 | 0·56 (0·29–1·11) | Date of analysis, smoking status, lifetime number of female sex partners, condom use in the past 3 months |
| Nielson 2009 | Cross-sectional | 13 types: 16, 18, 31, 33, 35, 39, 45, 51, 52, 56, 58, 59, 68 | 18–40 | 84·1 | 47·5 | Glans | 444 | 0·47 (0·22–0·99) | Date of analysis, smoking status, lifetime number of female sex partners, condom use in the past 3 months |
| Nielson 2009 | Cross-sectional | 13 types: 16, 18, 31, 33, 35, 39, 45, 51, 52, 56, 58, 59, 68 | 18–40 | 84·1 | 47·5 | Shaft | 449 | 0·50 (0·25–1·00) | Date of analysis, smoking status, lifetime number of female sex partners, condom use in the past 3 months |
| Shapiro 2022 | Cohort[b] | 22 types: 16, 18, 26, 31, 33, 34, 35, 39, 45, 51, 52, 53, 56, 58, 59, 66, 67, 68, 69, 70, 73, 82 | 17–45 | 47·2 | 54·6 | Combined | 413 | 0·87 (0·59–1·27) | Propensity score (male's and female's age, region born, race, educational attainment, smoking history, same-sex partner history, number of sexual partners, number of vaginal sexual partners, age at first sex; female's vaccination against HPV; couple's marital status, condom use, frequency of sex, concurrent partner at future visit) |
| Baldwin 2004 | Cross-sectional | 18 types: 16, 18, 26, 31, 33, 35, 39, 45, 51, 52, 55, 56, 58, 59, 68, 73, 82, 83 | 18–70 | 67·4 | 28·2 | Combined | 344 | 0·44 (0·22–0·90) | Sexual frequency per month, condom use in past 3 months |

| | | | | | | | | | |
|---|---|---|---|---|---|---|---|---|---|
| Hernandez 2008 | Cross-sectional | 19 types: 16, 18, 26, 31, 33, 35, 39, 45, 51–53, 56, 58, 59, 66, 68, 73, 82, 82/IS39 | NR | 82·6 | NR | Combined | 172 | 0·82 (0·28–2·38) | Age, birthplace, race/ethnicity, education level, lifetime number of female sex partners, history of sex with men, age at initial sex, condom use, history of genital warts, history of cigarette smoking |
| Hernandez 2008 | Cross-sectional | 19 types: 16, 18, 26, 31, 33, 35, 39, 45, 51–53, 56, 58, 59, 66, 68, 73, 82, 82/IS39 | NR | 82·6 | NR | Glans | 258 | 0·40 (0·18–0·90) | Age, birthplace, race/ethnicity, education level, lifetime number of female sex partners, history of sex with men, age at initial sex, condom use, history of genital warts, history of cigarette smoking |
| Hernandez 2008 | Cross-sectional | 19 types: 16, 18, 26, 31, 33, 35, 39, 45, 51–53, 56, 58, 59, 66, 68, 73, 82, 82/IS39 | NR | 82·6 | NR | Shaft | 243 | 0·70 (0·32–1·52) | Age, birthplace, race/ethnicity, education level, lifetime number of female sex partners, history of sex with men, age at initial sex, condom use, history of genital warts, history of cigarette smoking |
| Svare 2002 | Cross-sectional | 4 types: 16, 18, 31, 33 | 18–≥40 | 16·4 | 45 | Combined | 134 | 0·4 (0·08–1·7) | Age, lifetime sex partners |
| *Low-risk HPV* | | | | | | | | | |
| Albero 2013 | Cohort[b] | 24 types: 6, 11, 26, 40, 42, 53, 54, 55, 61, 62, 64, 66, 67, 69, 70, 71, 72, 73, 81, 82, 82/IS39, 83, 84, 89 | 18–70 | 35·9 | 66·7 | Combined | 3969 | 0·76 (0·67–0·87)[d] | None |
| Smith 2021 | RCT[c] | 22 types: 6, 11, 26, 34, 40, 42, 43, 44, 53, 54, 55, 57, 61, 70, 71, 72, 73, 81, 82/MM4, 82/IS39, 83, 84, 89[e] | 18–24 | 50·6 | 50[d] | Combined | 2193 | PR 0·59 (0·45–0·77) | None |
| Smith 2021 | RCT[c] | 22 types: 6, 11, 26, 34, 40, 42, 43, 44, 53, 54, 55, 57, 61, 70, 71, 72, 73, 81, 82/MM4, 82/IS39, 83, 84, 89[e] | 18–24 | 50·6 | 50[d] | Glans | 2193 | PRR 0·49 (0·36–0·65) | None |
| Smith 2021 | RCT[c] | 22 types: 6, 11, 26, 34, 40, 42, 43, 44, 53, 54, 55, 57, 61, 70, 71, 72, 73, 81, 82/MM4, 82/IS39, 83, 84, 89[e] | 18–24 | 50·6 | 50[d] | Shaft | 2193 | PRR 0·81 (0·54–1·19) | None |
| Tobian 2009 | RCT[c] | 23 types: 6, 11, 26, 40, 42, 53, 54, 55, 61, 62, 64, 67, 69, 70, 71, 72, 73, 81, 82, 82/IS39, 83, 84, 89 | 15–49 | 44·8 | 62·2 | Glans | 520 | RR 0·66 (0·49–0·91) | None |
| Nielson 2009 | Cross-sectional | 24 types: 6, 11, 26, 40, 42, 53, 54, 55, 61, 62, 64, 66, 67, 69, 70, 71, 72, 73, 81, 82, 82/IS39, 83, 84, 89 | 18–40 | 84·1 | 47·5 | Combined | 421 | 0·91 (0·47–1·78) | Date of analysis, smoking status, lifetime number of female sex partners, condom use in the past 3 months |

| | | | | | | | | | |
|---|---|---|---|---|---|---|---|---|---|
| Nielson 2009 | Cross-sectional | 24 types: 6, 11, 26, 40, 42, 53, 54, 55, 61, 62, 64, 66, 67, 69, 70, 71, 72, 73, 81, 82, 82/IS39, 83, 84, 89 | 18–40 | 84·1 | 47·5 | Glans | 444 | 0·62 (0·29–1·29) | Date of analysis, smoking status, lifetime number of female sex partners, condom use in the past 3 months |
| Nielson 2009 | Cross-sectional | 24 types: 6, 11, 26, 40, 42, 53, 54, 55, 61, 62, 64, 66, 67, 69, 70, 71, 72, 73, 81, 82, 82/IS39, 83, 84, 89 | 18–40 | 84·1 | 47·5 | Shaft | 449 | 0·85 (0·40–1·80) | Date of analysis, smoking status, lifetime number of female sex partners, condom use in the past 3 months |
| Shapiro 2022 | Cohort[b] | 14 types: 6, 11, 40, 42, 44, 54, 61, 62, 71, 72, 81, 83, 84, 89 | 17–45 | 47·2 | 54·6 | Combined | 413 | 0·90 (0·59–1·37) | Propensity score (male's and female's age, region born, race, educational attainment, smoking history, same-sex partner history, number of sexual partners, number of vaginal sexual partners, age at first sex; female's vaccination against HPV; couple's marital status, condom use, frequency of sex, concurrent partner at future visit) |
| Baldwin 2004 | Cross-sectional | 9 types: 6, 11, 40, 42, 53, 54, 57, 66, 84 | 18–70 | 67·4 | 28·2 | Glans | 344 | 0·44 (0·23–0·81) | Genital warts, condom use with last anal sex |
| Hernandez 2008 | Cross-sectional | 18 types: 6, 11, 40, 42, 54, 55, 61, 62, 64, 67, 69, 70, 71, 72, 81, 83, 84, 89 | NR | 80·9 | NR | Combined | 188 | 0·61 (0·25–1·47) | Age, birthplace, race/ethnicity, education level, lifetime number of female sex partners, history of sex with men, age at initial sex, condom use, history of genital warts, history of cigarette smoking |
| Hernandez 2008 | Cross-sectional | 18 types: 6, 11, 40, 42, 54, 55, 61, 62, 64, 67, 69, 70, 71, 72, 81, 83, 84, 89 | NR | 80·9 | NR | Glans | 280 | 0·52 (0·25–1·08) | Age, birthplace, race/ethnicity, education level, lifetime number of female sex partners, history of sex with men, age at initial sex, condom use, history of genital warts, history of cigarette smoking |
| Hernandez 2008 | Cross-sectional | 18 types: 6, 11, 40, 42, 54, 55, 61, 62, 64, 67, 69, 70, 71, 72, 81, 83, 84, 89 | NR | 80·9 | NR | Shaft | 292 | 0·59 (0·30–1·16) | Age, birthplace, race/ethnicity, education level, lifetime number of female sex partners, history of sex with men, age at initial sex, condom use, history of genital warts, history of cigarette smoking |
| Svare 2002 | Cross-sectional | 2 types: 6, 11 | 18–≥40 | 17·3 | 45 | Combined | 127 | 0·8 (0·1–4·1) | Age, number of sex partners in past year, ever had genital warts |

Abbreviations: CI, confidence interval; HPV, human papillomavirus; MC, male circumcision; NR, not reported; OR, odds ratio; PR, prevalence ratio; RCT, randomized controlled trial; RR, risk ratio

[a] All estimates presented are odds ratios, unless otherwise specified

[b] Cohort study analyzed cross-sectionally

[c] RCT analyzed cross-sectionally

[d] Calculation was performed manually
[e] Outcome was infection exclusively with low-risk types

**Supplementary Table 4: Studies of MC and HPV incidence in males by HPV risk grouping**

| First author (year) | Study design | HPV types detected | Age at baseline (range) | Circumcision prevalence (%) | HPV prevalence at baseline (%) | Sampling site | Number analyzed | Effect estimate: HR (95% CI)[a] | Covariate adjustment |
|---|---|---|---|---|---|---|---|---|---|
| *Any HPV* | | | | | | | | | |
| Albero 2014 | Cohort | 37 types: 16, 18, 31, 33, 35, 39, 45, 51, 52, 56, 58, 59, 68, 6, 11, 26, 40, 42, 53, 54, 55, 61, 62, 64, 66, 67, 69, 70, 71, 72, 73, 81, 82, 82/IS39, 83, 84, 89 | 18–70 | 36·4 | 66·8 | Combined | 4,033 | 1·08 (0·91–1·27) | Country, age, marital status, lifetime number of female sexual partners, recent number of female sexual partners, recent number of male anal sex partners, six-month visit compliance status |
| Smith 2021 | RCT | 44 types: 6, 11, 16, 18, 26, 30, 31, 32, 33, 34, 35, 39, 40, 42, 43, 44, 45, 51, 52, 53, 54, 55, 56, 57, 58, 59, 61, 64, 66, 67, 68, 69, 70, 71, 72, 73, 81, 82/MM4, 82/IS39, 83, 84, 85, 86, 89, JC9710 | 18–24 | 49·6 | 50[b] | Combined | 1,096 | 0·56 (0·45–0·70) | None |
| Smith 2021 | RCT | 44 types: 6, 11, 16, 18, 26, 30, 31, 32, 33, 34, 35, 39, 40, 42, 43, 44, 45, 51, 52, 53, 54, 55, 56, 57, 58, 59, 61, 64, 66, 67, 68, 69, 70, 71, 72, 73, 81, 82/MM4, 82/IS39, 83, 84, 85, 86, 89, JC9710 | 18–24 | 49·6 | 50[b] | Glans | 1,196 | 0·51 (0·43–0·61) | None |
| Smith 2021 | RCT | 44 types: 6, 11, 16, 18, 26, 30, 31, 32, 33, 34, 35, 39, 40, 42, 43, 44, 45, 51, 52, 53, 54, 55, 56, 57, 58, 59, 61, 64, 66, 67, 68, 69, 70, 71, 72, 73, 81, 82/MM4, 82/IS39, 83, 84, 85, 86, 89, JC9710 | 18–24 | 49·6 | 50[b] | Shaft | 1,795 | 1·01 (0·87–1·17) | None |
| Lu 2009 | Cohort | 37 types: 16, 18, 31, 33, 35, 39, 45, 51, 52, 56, 58, 59, 68, 6, 11, 26, 40, 42, 53, 54, 55, 61, 62, 64, 66, 67, 69, 70, 71, 72, 73, 81, 82, 82/IS39, 83, 84, 89 | 18–44 | 87·7 | NR | Combined | 285 | 0·8 (0·4–1·9) | Cigarette smoking, lifetime number of sexual partners |
| Partridge 2007 | Cohort | 37 types: 16, 18, 26, 31, 33, 35, 39, 45, 51, 52, 53, 56, 58, 59, 66, 67, 68, 73, 82, 6, 11, 40, 42, 54, 55, 57, 61, 62, 64, 69, 70, 71, 72, 81, 83, 84, 89 | 18–20 | 76·7 | 25·8 | Combined | 240 | 1·1 (0·6–2·0) | None |

| | | | | | | | | | |
|---|---|---|---|---|---|---|---|---|---|
| Lajous 2005 | Cohort | 27 types: 16, 18, 31, 33, 35, 39, 45, 51, 52, 53, 56, 58, 59, 68, 73, 82, 6, 11, 26, 40, 42, 54, 55, 57, 66, 83, 84 | 16–40 | 16·7 | 44·6 | Combined | 210 | OR 1·12 (0·45–2·80) | Age, SES, lifetime number of partners |
| Shapiro 2022 | Cohort | 36 types: 6, 11, 16, 18, 26, 31, 33, 34, 35, 39, 40, 42, 44, 45, 51, 52, 53, 54, 56, 58, 59, 61, 62, 66, 67, 68, 69, 70, 71, 72, 73, 81, 82, 83, 84, 89 | 17–45 | 47·2 | 54·6 | Combined | 413 | IRR 0·77 (0·37–1·60) | Propensity score (male's and female's age, region born, race, educational attainment, smoking history, same-sex partner history, number of sexual partners, number of vaginal sexual partners, age at first sex; female's vaccination against HPV; couple's marital status, condom use, frequency of sex, concurrent partner at future visit) |
| *High-risk HPV* | | | | | | | | | |
| Albero 2014 | Cohort | 13 types: 16, 18, 31, 33, 35, 39, 45, 51, 52, 56, 58, 59, 68 | 18–70 | 36·4 | 66·8 | Combined | 4,033 | 1·11 (0·94–1·31) | Country, age, marital status, lifetime number of female sexual partners, recent number of female sexual partners, and recent number of male anal sex partners |
| Smith 2021 | RCT | 14 types: 16, 18, 31, 33, 35, 39, 45, 51, 52, 56, 58, 59, 66, 68 | 18–24 | 50·1 | 50[b] | Combined | 1,335 | 0·58 (0·49–0·69) | None |
| Smith 2021 | RCT | 14 types: 16, 18, 31, 33, 35, 39, 45, 51, 52, 56, 58, 59, 66, 68 | 18–24 | 50·1 | 50[b] | Glans | 1,442 | 0·48 (0·40–0·57) | None |
| Smith 2021 | RCT | 14 types: 16, 18, 31, 33, 35, 39, 45, 51, 52, 56, 58, 59, 66, 68 | 18–24 | 50·1 | 50[b] | Shaft | 1891 | 0·98 (0·82–1·17) | None |
| Tobian 2012 | RCT | 14 types: 6, 18, 31, 33, 35, 39, 45, 51, 52, 56, 58, 59, 66, 68 | 15–49 | NR | NR | Glans | 776 | IRR 0·70 (0·55–0·89) | Age, marital status, non-marital relationships, number of sexual partners during past year, condom use past year, self-reported urethral discharge |
| Vanbuskirk 2011 | Cohort | 19 types: 16, 18, 26, 31, 33, 35, 39, 45, 51, 52, 53, 56, 58, 59, 66, 68, 73, 82, 82/IS39 | 18–20 | 75·3 | 20 | Combined | 477 | 1·1 (0·8–1·4) | None |
| Gray 2010 | RCT | 14 types: 16, 18, 31, 33, 35, 39, 45, 51, 52, 56, 58, 59, 66, 68 | 15–49 | 46·4 | 38·9 | Glans | 448 | IRR 0·67 (0·50–0·91) | Age, education, condom use, alcohol consumption with sex, number of sex partners |
| Lu 2009 | Cohort | 13 types: 16, 18, 31, 33, 35, 39, 45, 51, 52, 56, 58, 59, 68 | 18–44 | 87·7 | NR | Combined | 285 | 1·7 (0·6–4·9) | Age at first sexual intercourse, lifetime number of sexual partners |

| | | | | | | | | | |
|---|---|---|---|---|---|---|---|---|---|
| Shapiro 2022 | Cohort | 22 types: 16, 18, 26, 31, 33, 34, 35, 39, 45, 51, 52, 53, 56, 58, 59, 66, 67, 68, 69, 70, 73, 82 | 17–45 | 47·2 | 54·6 | Combined | 19 | IRR 1·19 (0·44–3·23) | Propensity score (male's and female's age, region born, race, educational attainment, smoking history, same-sex partner history, number of sexual partners, number of vaginal sexual partners, age at first sex; female's vaccination against HPV; couple's marital status, condom use, frequency of sex, concurrent partner at future visit) |
| *Low-risk HPV* | | | | | | | | | |
| Albero 2014 | Cohort | 24 types: 6, 11, 26, 40, 42, 53, 54, 55, 61, 62, 64, 66, 67, 69, 70, 71, 72, 73, 81, 82, 82/IS39, 83, 84, 89 | 18–70 | 36·4 | 66·8 | Combined | 4,033 | 1·11 (0·94–1·30) | Country, age, marital status, lifetime number of female sexual partners, recent number of female sexual partners, lifetime number of male anal sex partners, and six-month visit compliance status |
| Smith 2021 | RCT | 22 types: 6, 11, 26, 34, 40, 42, 43, 44, 53, 54, 55, 57, 61, 70, 71, 72, 73, 81, 82/MM4, 82/IS39, 83, 84, 89[c] | 18–24 | 50·0[a] | 50[b] | Combined | 1,851 | 0·61 (0·51–0·73) | None |
| Smith 2021 | RCT | 22 types: 6, 11, 26, 34, 40, 42, 43, 44, 53, 54, 55, 57, 61, 70, 71, 72, 73, 81, 82/MM4, 82/IS39, 83, 84, 89[c] | 18–24 | 50·0[a] | 50[b] | Glans | 1,863 | 0·54 (0·44–0·65) | None |
| Smith 2021 | RCT | 22 types: 6, 11, 26, 34, 40, 42, 43, 44, 53, 54, 55, 57, 61, 70, 71, 72, 73, 81, 82/MM4, 82/IS39, 83, 84, 89[c] | 18–24 | 50·0[a] | 50[b] | Shaft | 2,048 | 0·86 (0·68–1·07) | None |
| Gray 2010 | RCT | 20 types: 6, 11, 26, 40, 42, 43, 53, 54, 55, 61, 67, 70, 71, 72, 73, 81, 82, 83, 84, 108 | 15–49 | 46·4 | 38·9 | Glans | 448 | IRR 0·84 (0·66–1·10) | None |
| Lu 2009[‡] | Cohort | 24 types: 6, 11, 26, 40, 42, 53, 54, 55, 61, 62, 64, 66, 67, 69, 70, 71, 72, 73, 81, 82, 82/IS39, 83, 84, 89 | 18–44 | 87·7 | NR | Combined | 285 | 1·0 (0·4–2·5) | None |
| Shapiro 2022 | Cohort | 14 types: 6, 11, 40, 42, 44, 54, 61, 62, 71, 72, 81, 83, 84, 89 | 17–45 | 47·2 | 54·6 | Combined | 42 | IRR 0·72 (0·21–2·45) | Propensity score (male's and female's age, region born, race, educational attainment, smoking history, same-sex partner history, number of sexual partners, number of vaginal sexual partners, age at |

| | | | | | | | | | |
|---|---|---|---|---|---|---|---|---|---|
| | | | | | | | | | first sex; female's vaccination against HPV; couple's marital status, condom use, frequency of sex, concurrent partner at future visit) |

Abbreviations: CI, confidence interval; HPV, human papillomavirus; HR, hazard ratio; IRR, incidence rate ratio; MC, male circumcision; NR, not reported; RCT, randomized controlled trial

[a] All estimates presented are hazard ratios, unless otherwise specified

[a] Calculation was performed manually

[b] Outcome was infection with exclusively low-risk types

#### Supplementary Table 5: Studies of MC and HPV clearance in males by HPV risk grouping

| First author (year) | Study design | HPV types detected | Age at baseline, years (range) | Circumcision prevalence (%) | HPV prevalence at baseline (%) | Sampling site | Number analyzed | Effect estimate: HR (95% CI)[a] | Covariate adjustment |
|---|---|---|---|---|---|---|---|---|---|
| *Any HPV* | | | | | | | | | |
| Albero 2014 | Cohort | 37 types: 16, 18, 31, 33, 35, 39, 45, 51, 52, 56, 58, 59, 68, 6, 11, 26, 40, 42, 53, 54, 55, 61, 62, 64, 66, 67, 69, 70, 71, 72, 73, 81, 82, 82/IS39, 83, 84, 89 | 18–70 | 36·4 | 66·8 | Combined | 4,033 | 0·95 (0·88–1·02) | Country, age, lifetime number of female sexual partners, recent number of male anal sex partners, smoking status, HPV status at baseline, six-month visit compliance status |
| Smith 2021 | RCT | 44 types: 6, 11, 16, 18, 26, 30, 31, 32, 33, 34, 35, 39, 40, 42, 43, 44, 45, 51, 52, 53, 54, 55, 56, 57, 58, 59, 61, 64, 66, 67, 68, 69, 70, 71, 72, 73, 81, 82/MM4, 82/IS39, 83, 84, 85, 86, 89, JC9710 | 18–24 | 49·8 | 50 | Combined | 2,331 | 1·98 (1·48–2·66) | None |
| Smith 2021 | RCT | 44 types: 6, 11, 16, 18, 26, 30, 31, 32, 33, 34, 35, 39, 40, 42, 43, 44, 45, 51, 52, 53, 54, 55, 56, 57, 58, 59, 61, 64, 66, 67, 68, 69, 70, 71, 72, 73, 81, 82/MM4, 82/IS39, 83, 84, 85, 86, 89, JC9710 | 18–24 | 49·8 | 50 | Glans | 2,032 | 1·90 (1·49–2·42) | None |
| Smith 2021 | RCT | 44 types: 6, 11, 16, 18, 26, 30, 31, 32, 33, 34, 35, 39, 40, 42, 43, 44, 45, 51, 52, 53, 54, 55, 56, 57, 58, 59, 61, 64, 66, 67, 68, 69, 70, 71, 72, 73, 81, 82/MM4, 82/IS39, 83, 84, 85, 86, 89, JC9710 | 18–24 | 49·8 | 50 | Shaft | 624 | 2·19 (1·34–3·58) | None |
| Hernandez 2010 | Cohort | 37 types: 16, 18, 31, 33, 35, 39, 45, 51, 52, 56, 58, 59, 68, 6, 11, 26, 40, 42, 53, 54, 55, 61, 62, 64, 66, 67, 69, 70, 71, 72, 73, 81, 82, 82/IS39, 83, 84, 89 | 18–79 | 81·2 | 50 | Combined | 357 | 0·96 (0·71–1·32) | Age, race/ethnicity, birthplace, education, lifetime number of female partners, history of sex with men, condom use during prior 4 months, history of genital warts |
| Hernandez 2010 | Cohort | 37 types: 16, 18, 31, 33, 35, 39, 45, 51, 52, 56, 58, 59, 68, 6, 11, 26, 40, 42, 53, 54, 55, 61, 62, 64, 66, 67, 69, 70, 71, 72, 73, 81, 82, 82/IS39, 83, 84, 89 | 18–79 | 81·2 | 50 | Glans | 357 | 1·69 (1·02–2·78) | Age, race/ethnicity, birthplace, education, lifetime number of female partners, history of sex with men, condom use during prior 4 months, history of genital warts |
| Hernandez 2010 | Cohort | 37 types: 16, 18, 31, 33, 35, 39, 45, 51, 52, 56, 58, 59, 68, 6, 11, 26, 40, 42, 53, 54, 55, 61, 62, 64, 66, 67, 69, 70, 71, 72, 73, 81, 82, 82/IS39, 83, 84, 89 | 18–79 | 81·2 | 50 | Shaft | 357 | 0·94 (0·63–1·41) | Age, race/ethnicity, birthplace, education, lifetime number of female partners, history of sex with men, condom use during prior 4 months, history of genital warts |

| | | | | | | | | | |
|---|---|---|---|---|---|---|---|---|---|
| Lu 2009 | Cohort | 37 types: 16, 18, 31, 33, 35, 39, 45, 51, 52, 56, 58, 59, 68, 6, 11, 26, 40, 42, 53, 54, 55, 61, 62, 64, 66, 67, 69, 70, 71, 72, 73, 81, 82, 82/IS39, 83, 84, 89 | 18–44 | 87·7 | NR | | 285 | 3·1 (1·2–8·2) | Cigarette smoking, lifetime number of sexual partners |
| Shapiro 2022 | Cohort | 36 types: 6, 11, 16, 18, 26, 31, 33, 34, 35, 39, 40, 42, 44, 45, 51, 52, 53, 54, 56, 58, 59, 61, 62, 66, 67, 68, 69, 70, 71, 72, 73, 81, 82, 83, 84, 89 | 17–45 | 47·2 | 54·6 | Combined | 131 | CRR 0·81 (0·52–1·24) | Propensity score (male's and female's age, region born, race, educational attainment, smoking history, same-sex partner history, number of sexual partners, number of vaginal sexual partners, age at first sex; female's vaccination against HPV; couple's marital status, condom use, frequency of sex, concurrent partner at future visit) |
| *High-risk HPV* | | | | | | | | | |
| Albero 2014 | Cohort | 13 types: 16, 18, 31, 33, 35, 39, 45, 51, 52, 56, 58, 59, 68 | 18–70 | 36·4 | 66·8 | Combined | 4,033 | 0·9 (0·81–1·00) | Country, age, lifetime number of female sexual partners, lifetime number of male anal sex partners, HPV status at baseline, six-month visit compliance status |
| Smith 2021 | RCT | 14 types: 16, 18, 31, 33, 35, 39, 45, 51, 52, 56, 58, 59, 66, 68 | 18–24 | 48·9 | 50 | Combined | 1,239 | 1·76 (1·29–2·39) | None |
| Smith 2021 | RCT | 14 types: 16, 18, 31, 33, 35, 39, 45, 51, 52, 56, 58, 59, 66, 68 | 18–24 | 48·9 | 50 | Glans | 1,051 | 1·73 (1·27–2·37) | None |
| Smith 2021 | RCT | 14 types: 16, 18, 31, 33, 35, 39, 45, 51, 52, 56, 58, 59, 66, 68 | 18–24 | 48·9 | 50 | Shaft | 356 | 1·15 (0·59–2·25) | None |
| Tobian 2012 | RCT | 14 types: 6, 18, 31, 33, 35, 39, 45, 51, 52, 56, 58, 59, 66, 68 | 15–49 | NR | NR | Shaft | 776 | RR 1·48 (1·26–1·74) | Age, occupation, marital status, self-reported urethral discharge, self-reported dysuria, enrollment syphilis status, HSV-2 status |
| Gray 2010 | RCT | 14 types: 16, 18, 31, 33, 35, 39, 45, 51, 52, 56, 58, 59, 66, 68 | 15–49 | 46·4 | 38·9 | Glans | 448 | CRR 1·39 (1·17–1·64) | Age, education, number of sex partners, condom use |
| Hernandez 2010 | Cohort | 13 types: 16, 18, 31, 33, 35, 39, 45, 51, 52, 56, 58, 59, 68 | 18–79 | 81·2 | 50 | Combined | 357 | 1·11 (0·60–2·08) | Age, race/ethnicity, birthplace, education, lifetime number of female partners, history of sex with men, condom use during prior 4 months, history of genital warts |
| Hernandez 2010 | Cohort | 13 types: 16, 18, 31, 33, 35, 39, 45, 51, 52, 56, 58, 59, 68 | 18–79 | 81·2 | 50 | Glans | 357 | 2·78 (1·10–7·14) | Age, race/ethnicity, birthplace, education, lifetime number of female partners, history of sex with men, condom use during prior 4 months, history of genital warts |

| | | | | | | | | | |
|---|---|---|---|---|---|---|---|---|---|
| Hernandez 2010 | Cohort | 13 types: 16, 18, 31, 33, 35, 39, 45, 51, 52, 56, 58, 59, 68 | 18–79 | 81·2 | 50 | Shaft | 357 | 1·67 (0·67–4·17) | Age, race/ethnicity, birthplace, education, lifetime number of female partners, history of sex with men, condom use during prior 4 months, history of genital warts |
| Lu 2009 | Cohort | 13 types: 16, 18, 31, 33, 35, 39, 45, 51, 52, 56, 58, 59, 68 | 18–44 | 87·7 | NR | Combined | 285 | 6·5 (2·1–19·7) | Age at first sexual intercourse, lifetime number of sexual partners |
| *Low-risk HPV* | | | | | | | | | |
| Albero 2014 | Cohort | 24 types: 6, 11, 26, 40, 42, 53, 54, 55, 61, 62, 64, 66, 67, 69, 70, 71, 72, 73, 81, 82, 82/IS39, 83, 84, 89 | 18–70 | 36·4 | 66·8 | Combined | 4,033 | 0·98 (0·89–1·07) | Country, age, recent number of female sexual partners, recent number of male anal sex partners, smoking status, HPV status at baseline, six-month visit compliance status |
| Smith 2021 | RCT | 22 types: 6, 11, 26, 34, 40, 42, 43, 44, 53, 54, 55, 57, 61, 70, 71, 72, 73, 81, 82/MM4, 82/IS39, 83, 84, 89[c] | 18–24 | 50[b] | 50 | Combined | 1,094 | 1·56 (1·21–2·00) | None |
| Smith 2021 | RCT | 22 types: 6, 11, 26, 34, 40, 42, 43, 44, 53, 54, 55, 57, 61, 70, 71, 72, 73, 81, 82/MM4, 82/IS39, 83, 84, 89[c] | 18–24 | 50[b] | 50 | Glans | 981 | 1·54 (1·29–1·85) | None |
| Smith 2021 | RCT | 22 types: 6, 11, 26, 34, 40, 42, 43, 44, 53, 54, 55, 57, 61, 70, 71, 72, 73, 81, 82/MM4, 82/IS39, 83, 84, 89[c] | 18–24 | 50[b] | 50 | Shaft | 268 | 1·31 (0·79–2·18) | None |
| Hernandez 2010 | Cohort | 24 types: 6, 11, 26, 40, 42, 53, 54, 55, 61, 62, 64, 66, 67, 69, 70, 71, 72, 73, 81, 82, 82/IS39, 83, 84, 89[c] | 18–79 | 81·2 | 50 | Combined | 357 | 0·87 (0·50–1·54) | Age, race/ethnicity, birthplace, education, lifetime number of female partners, history of sex with men, condom use during prior 4 months, history of genital warts |
| Hernandez 2010 | Cohort | 24 types: 6, 11, 26, 40, 42, 53, 54, 55, 61, 62, 64, 66, 67, 69, 70, 71, 72, 73, 81, 82, 82/IS39, 83, 84, 89[c] | 18–79 | 81·2 | 50 | Glans | 357 | 2·00 (1·02–4·00) | Age, race/ethnicity, birthplace, education, lifetime number of female partners, history of sex with men, condom use during prior 4 months, history of genital warts |
| Hernandez 2010 | Cohort | 24 types: 6, 11, 26, 40, 42, 53, 54, 55, 61, 62, 64, 66, 67, 69, 70, 71, 72, 73, 81, 82, 82/IS39, 83, 84, 89[c] | 18–79 | 81·2 | 50 | Shaft | 357 | 1·16 (0·56–2·44) | Age, race/ethnicity, birthplace, education, lifetime number of female partners, history of sex with men, condom use during prior 4 months, history of genital warts |
| Lu 2009 | Cohort | 24 types: 6, 11, 26, 40, 42, 53, 54, 55, 61, 62, 64, 66, 67, 69, 70, 71, 72, 73, 81, 82, 82/IS39, 83, 84, 89[c] | 18–44 | 87·7 | NR | Combined | 285 | 1·6 (0·7–3·7) | None |

Abbreviations: CI, confidence interval; CRR, clearance rate ratio; HPV, human papillomavirus; HR, hazard ratio; MC, male circumcision; NR, not reported; RCT, randomized controlled trial; RR, risk ratio

[a] All estimates presented are hazard ratios, unless otherwise specified

[b] Calculation was performed manually

[c] Outcome was infection with exclusively low-risk type(s)

**Supplementary Table 6: Studies of MC and various HPV infection outcomes in females by HPV risk grouping**

| First author (year) | Study design | HPV types detected | Age at baseline, years (range) | Circumcision prevalence (%) | HPV prevalence at baseline (%) | Number analyzed | Effect estimate: OR (95% CI) | Covariate adjustment |
|---|---|---|---|---|---|---|---|---|
| *Prevalence, any HPV* | | | | | | | | |
| Roura 2012 | Cross-sectional | 27 types: 16, 18, 26, 31, 33, 35, 39, 45, 51, 52, 53, 56, 58, 59, 66, 68, 73, 82, 6, 11, 40, 43, 44, 54, 69/71, 70, 74) | 18–65 | 13·5 | 19·3 | 2,735 | 0·8 (0·6–1·1) | Age, autonomous community, country of birth, marital status, level of education, smoking habits, lifetime number of sexual partners, history of genital warts |
| Wawer 2011 | RCT[b] | 27 types: 16, 18, 26, 31, 33, 35, 39, 45, 51, 52, 55, 56, 58, 59, 66, 68, 6, 11, 26, 40, 42, 53, 54, 55, 57, 73, 82, 83, 84) | 15–49 | 52·7 | 55·8[d] | 1,032 | **PR 0·81 (0·72–0·92)** | None |
| Shapiro 2022 | Cohort[c] | 36 types: 6, 11, 16, 18, 26, 31, 33, 34, 35, 39, 40, 42, 44, 45, 51, 52, 53, 54, 56, 58, 59, 61, 62, 66, 67, 68, 69, 70, 71, 72, 73, 81, 82, 83, 84, 89 | 18–25 | 47·2 | 54·6 | 413 | 1·05 (0·75–1·46) | Propensity score (male's and female's age, region born, race, educational attainment, smoking history, same-sex partner history, number of sexual partners, number of vaginal sexual partners, age at first sex; female's vaccination against HPV; couple's marital status, condom use, frequency of sex, concurrent partner at future visit) |
| Mbulawa 2009 | Cross-sectional | 37 types: 16, 18, 31, 33, 35, 39, 45, 51, 52, 56, 58, 59, 68, 73, 82, 26, 53, 66, 6, 11, 40, 42, 54, 55, 61, 62, 64, 67, 69, 70, 71, 72, 81, 82/IS39, 83, 84, 89 | 18–65 | 92·1 | 31 | 202 | 0·42 (0·14–1·20)[d] | None |
| Contreras 2008 | Cross-sectional | 14+ types: 5, 6, 8, 11, 16, 18, 31, 33, 35, 39, 45, 51, 56, 58, others | 18–55 | 19·7 | 30 | 61 | **9 (1·2–64·4)** | Age, having more than 1 sexual partner |
| *Prevalence, high-risk HPV* | | | | | | | | |
| Roura 2012 | Cross-sectional | 18 types: 16, 18, 26, 31, 33, 35, 39, 45, 51, 52, 53, 56, 58, 59, 66, 68, 73, 82 | 18–65 | 13·5 | 19·3 | 2,735 | 0·7 (0·5–1·0) | Age, autonomous community, country of birth, marital status, level of education, smoking habits, lifetime number of sexual partners, history of genital warts |
| Wawer 2011 | RCT[b] | 14 types: 16, 18, 26, 31, 33, 35, 39, 45, 51, 52, 55, 56, 58, 59, 66, 68 | 15–49 | 52·7 | 55·8[d] | 1,032 | **PR 0·72 (0·60–0·85)** | None |
| Shapiro 2022 | Cohort[c] | 22 types: 16, 18, 26, 31, 33, 34, 35, 39, 45, 51, 52, 53, 56, 58, 59, 66, 67, 68, 69, 70, 73, 82 | 18–25 | 47·2 | 54·6 | 413 | 1·06 (0·74–1·52) | Propensity score (male's and female's age, region born, race, educational attainment, smoking history, same-sex partner history, number of sexual partners, number of vaginal sexual partners, age at first sex; female's vaccination against HPV; couple's |

| | | | | | | | | |
|---|---|---|---|---|---|---|---|---|
| | | | | | | | | marital status, condom use, frequency of sex, concurrent partner at future visit) |
| Obiri-Yeboah 2017 | Cross-sectional | 19 types: 16, 18, 26, 31, 33, 35, 39, 45, 51, 52, 53, 56, 58, 59, 66, 68, 69, 73, 82 | 18-NR | 93·5 | 42·6 | 170 | **RR 0·5 (0·3–0·9)** | None |
| *Prevalence, low-risk HPV* | | | | | | | | |
| Roura 2012 | Cross-sectional | 9 types: 6, 11, 40, 43, 44, 54, 69/71, 70, 74 | 18–65 | 13·5 | 19·3 | 2,735 | 0·7 (0·5–1·0) | Age, autonomous community, country of birth, marital status, level of education, smoking habits, lifetime number of sexual partners, history of genital warts |
| Wawer 2011 | RCT[b] | 13 types: 6, 11, 26, 40, 42, 53, 54, 55, 57, 73, 82, 83, 84 | 15–49 | 52·7 | 55·8[d] | 1,032 | **PR 0·77 (0·66–0·90)** | None |
| Shapiro 2022 | Cohort[c] | 14 types: 6, 11, 40, 42, 44, 54, 61, 62, 71, 72, 81, 83, 84, 89 | 18–25 | 47·2 | 54·6 | 413 | 1·07 (0·72–1·60) | Propensity score (male's and female's age, region born, race, educational attainment, smoking history, same-sex partner history, number of sexual partners, number of vaginal sexual partners, age at first sex; female's vaccination against HPV; couple's marital status, condom use, frequency of sex, concurrent partner at future visit) |
| *Incidence, any HPV* | | | | | | | | |
| Shapiro 2022 | Cohort | 36 types: 6, 11, 16, 18, 26, 31, 33, 34, 35, 39, 40, 42, 44, 45, 51, 52, 53, 54, 56, 58, 59, 61, 62, 66, 67, 68, 69, 70, 71, 72, 73, 81, 82, 83, 84, 89 | 18–25 | 47·2 | 54·6 | 61 | IRR 0·59 (0·16–2·20) | Propensity score (male's and female's age, region born, race, educational attainment, smoking history, same-sex partner history, number of sexual partners, number of vaginal sexual partners, age at first sex; female's vaccination against HPV; couple's marital status, condom use, frequency of sex, concurrent partner at future visit) |
| *Incidence, high-risk HPV* | | | | | | | | |
| Wawer 2011 | RCT | 14 types: 16, 18, 26, 31, 33, 35, 39, 45, 51, 52, 55, 56, 58, 59, 66, 68 | 15–49 | 52·2 | 55·8[d] | 1,051 | **IRR 0·77 (0·63–0·93)** | None |
| Shapiro 2022 | Cohort | 22 types: 16, 18, 26, 31, 33, 34, 35, 39, 45, 51, 52, 53, 56, 58, 59, 66, 67, 68, 69, 70, 73, 82 | 18–25 | 47·2 | 54·6 | 32 | IRR 0·80 (0·12–5·38) | Propensity score (male's and female's age, region born, race, educational attainment, smoking history, same-sex partner history, number of sexual partners, number of vaginal sexual partners, age at first sex; female's vaccination against HPV; couple's marital status, condom use, frequency of sex, concurrent partner at future visit) |
| *Incidence, low-risk HPV* | | | | | | | | |

| | | | | | | | | |
|---|---|---|---|---|---|---|---|---|
| Wawer 2011 | RCT | 13 types: 6, 11, 26, 40, 42, 53, 54, 55, 57, 73, 82, 83, 84 | 15–49 | 52·2 | 55·8[d] | 1,051 | IRR 0·83 (0·69–1·00) | None |
| Shapiro 2022 | Cohort | 14 types: 6, 11, 40, 42, 44, 54, 61, 62, 71, 72, 81, 83, 84, 89 | 18–25 | 47·2 | 54·6 | 30 | IRR 0·23 (0·03–1·55) | Propensity score (male's and female's age, region born, race, educational attainment, smoking history, same-sex partner history, number of sexual partners, number of vaginal sexual partners, age at first sex; female's vaccination against HPV; couple's marital status, condom use, frequency of sex, concurrent partner at future visit) |
| *Clearance, high-risk HPV* | | | | | | | | |
| Wawer 2011 | RCT | 14 types: 16, 18, 26, 31, 33, 35, 39, 45, 51, 52, 55, 56, 58, 59, 66, 68 | 15–49 | 52·2 | 55·8[d] | 1,051 | **RR 1·12 (1·02–1·22)** | None |

Abbreviations: CI, confidence interval; HPV, human papillomavirus; IRR, incidence rate ratio; MC, male circumcision; NR, not reported; OR, odds ratio; RCT, randomized controlled trial; RR, risk ratio

[a] All estimates presented are odds ratios, unless otherwise specified

[b] RCT analyzed cross-sectionally

[c] Cohort study analyzed cross-sectionally

[d] Calculation was performed manually

**Supplementary Table 7: Risk of bias assessment, cross-sectional studies**

| First author & year | Sample representativeness (*) | Sample description | Justification for sample size (*) | Comparability of respondents and non-respondents (*) | Ascertainment of exposure status (**, *) | Comparability of subjects in outcome groups (**, *) | Ascertainment of outcome status (**, *) | Appropriateness of statistical test (*) | Total score out of 10 |
|---|---|---|---|---|---|---|---|---|---|
| Albero 2013 | Truly representative (*) | Males from the general population, universities, and organized healthcare systems | Not justified | No description of the response rate or the characteristics of the responders and the non-responders | Clinician assessment or validation (**) | Adjusts for or restricts to at least 2 key variables (**) | Laboratory test (**) | Described and appropriate (*) | **8** |
| Baldwin 2004 | Selected group of users | Males from an STI clinic | Not justified | The response rate is unsatisfactory, or the comparability between respondents and non-respondents is unsatisfactory | Clinician assessment or validation (**) | Adjusts for or restricts to at least 2 key variables (**) | Laboratory test (**) | Described and appropriate (*) | **7** |
| Bleeker 2005 | Selected group of users | Males from a non-STI dermatology clinic and male partners of females with CIN | Not justified | Comparability between respondents and non-respondents characteristics is established, and the response rate is satisfactory (*) | Clinician assessment or validation (**) | Does not restrict to or adjust for any variables | Laboratory test (**) | Described and appropriate (*) | **6** |
| Castellsagué 2002 | Selected group of users | Participants from one of seven case-control studies of cervical cancer | Not justified | The response rate is unsatisfactory, or the comparability between respondents and non-respondents is unsatisfactory | Clinician assessment or validation (**) | Adjusts for or restricts to at least 2 key variables (**) | Laboratory test (**) | Described and appropriate (*) | **7** |
| Contreras 2008 | Selected group of users | Females with rheumatoid arthritis | Justified and satisfactory (*) | No description of the response rate or the characteristics of the responders and the non-responders | Self-report or partner report (*) | Adjusts for or restricts to at least 2 key variables (**) | Laboratory test (**) | Described and appropriate (*) | **7** |
| Da Rocha 2015 | Somewhat representative (*) | Males from an STI clinic, dermatology clinic, university, and metallurgical factory | Justified and satisfactory (*) | No description of the response rate or the characteristics of the responders and the non-responders | Self-report or partner report (*) | Does not restrict to or adjust for any variables | Laboratory test (**) | Described and appropriate (*) | **6** |
| Hebnes 2021 | Somewhat representative (*) | Males from the military | Not justified | No description of the response rate or the characteristics of the responders | Clinician assessment or validation (**) | Adjusts for or restricts to at least 2 key variables (**) | Laboratory test (**) | Described and appropriate (*) | **8** |

| | | | | | | | | | |
|---|---|---|---|---|---|---|---|---|---|
| | | | | and the non-responders | | | | | |
| Hernandez 2008 | Somewhat representative (*) | Male university students | Not justified | No description of the response rate or the characteristics of the responders and the non-responders | Clinician assessment or validation (**) | Adjusts for or restricts to at least 2 key variables (**) | Laboratory test (**) | Described and appropriate (*) | **8** |
| Mbulawa 2008 | Selected group of users | Couples who were ineligible for or had completed an HIV transmission trial | Not justified | No description of the response rate or the characteristics of the responders and the non-responders | Self-report or partner report (*) | Does not restrict to or adjust for any variables | Laboratory test (**) | Described and appropriate (*) | **4** |
| Nielson 2009 | Truly representative (*) | Males from the general population | Not justified | The response rate is unsatisfactory, or the comparability between respondents and non-respondents is unsatisfactory | Self-report or partner report (*) | Adjusts for or restricts to at least 2 key variables (**) | Laboratory test (**) | Described and appropriate (*) | **7** |
| Obiri-Yeboah 2017 | Somewhat representative (*) | Females attending a hospital HIV or medical outpatient clinic | Justified and satisfactory (*) | No description of the response rate or the characteristics of the responders and the non-responders | Self-report or partner report (*) | Does not restrict to or adjust for any variables | Laboratory test (**) | Described and appropriate (*) | **6** |
| Ogilvie 2009 | Selected group of users | Males from an STI clinic | Not justified | No description of the response rate or the characteristics of the responders and the non-responders | Clinician assessment or validation (**) | Does not restrict to or adjust for any variables | Laboratory test (**) | Described and appropriate (*) | **5** |
| Olesen 2019 | Truly representative (*) | Males from urban and rural areas | Not justified | No description of the response rate or the characteristics of the responders and the non-responders | Clinician assessment or validation (**) | Does not restrict to or adjust for any variables | Laboratory test (**) | Described and appropriate (*) | **6** |
| Rocha 2012 | Selected group of users | Heterosexual couples in which the female partner had HPV-related cervical lesions | Not justified | No description of the response rate or the characteristics of the responders and the non-responders | Clinician assessment or validation (**) | Does not restrict to or adjust for any variables | Laboratory test (**) | Described and appropriate (*) | **5** |
| Rombaldi 2006 | Selected group of users | Male sexual partners of females with CIN | Not justified | No description of the response rate or the characteristics of the responders | Self-report or partner report (*) | Does not restrict to or adjust for any variables | Laboratory test (**) | Described and appropriate (*) | **4** |

| | | | | | | | | | |
|---|---|---|---|---|---|---|---|---|---|
| | | | | and the non-responders | | | | | |
| Roura 2012 | Somewhat representative (*) | Females attending routine cervical cancer screening | Justified and satisfactory (*) | The response rate is unsatisfactory, or the comparability between respondents and non-respondents is unsatisfactory | Self-report or partner report (*) | Adjusts for or restricts to at least 2 key variables (**) | Laboratory test (**) | Described and appropriate (*) | **8** |
| Shin 2004 | Somewhat representative (*) | Male university students | Not justified | Comparability between respondents and non-respondents characteristics is established, and the response rate is satisfactory (*) | Self-report or partner report (*) | Adjusts for or restricts to at least 2 key variables (**) | Laboratory test (**) | Described and appropriate (*) | **8** |
| Svare 2002 | Selected group of users | Males attending STI clinic | Not justified | No description of the response rate or the characteristics of the responders and the non-responders | Self-report or partner report (*) | Adjusts for or restricts to at least 2 key variables (**) | Laboratory test (**) | Described and appropriate (*) | **6** |
| Vaccarella 2006 | Selected group of users | Males requesting a vasectomy | Not justified | Comparability between respondents and non-respondents characteristics is established, and the response rate is satisfactory (*) | Clinician assessment or validation (**) | Adjusts for or restricts to at least 2 key variables (**) | Laboratory test (**) | Described and appropriate (*) | **8** |
| Vardas 2011 | Somewhat representative (*) | Heterosexual males with 1–5 female lifetime sexual partners | Not justified | No description of the response rate or the characteristics of the responders and the non-responders | Clinician assessment or validation (**) | Adjusts for or restricts to at least 2 key variables (**) | Laboratory test (**) | Described and appropriate (*) | **8** |

**Supplementary Table 8: Risk of bias assessment, cohort studies**

| First author & year | Representativeness of exposed cohort (*) | Description of cohort | Selection of non-exposed cohort (*) | Ascertainment of exposure status (*) | Demonstration of absence of outcome at beginning of study (*) | Comparability of cohorts on basis of design or analysis (**, *) | Ascertainment of outcome status (**, *) | Duration of follow-up (*) | Adequacy of follow-up (*) | Total score out of 10 |
|---|---|---|---|---|---|---|---|---|---|---|
| Albero 2014 | Truly representative (*) | Males from the general population, universities, and organized healthcare systems | Drawn from the same community as exposed cohort (*) | Clinician assessment (*) | Yes (*) | Restricts to or adjusts for at least 2 key variables (**) | Regular follow-up (*) | Mean or median follow-up of at least 3 months (*) | Not reported | **7** |
| Hernandez 2010 | Somewhat representative (*) | Male university students | Drawn from the same community as exposed cohort (*) | Clinician assessment (*) | Yes (*) | Restricts to or adjusts for at least 2 key variables (**) | Regular follow-up (*) | Mean or median follow-up of at least 3 months (*) | Not reported | **7** |
| Lajous 2005 | Selected group | Healthy military males | Drawn from the same community as exposed cohort (*) | Self-report or partner report | Yes (*) | Restricts to or adjusts for at least 2 key variables (**) | Regular follow-up (*) | Mean or median follow-up of at least 3 months (*) | Loss to follow-up may introduce bias | **5** |
| Lu 2009 | Truly representative (*) | Males from the general population | Drawn from the same community as exposed cohort (*) | Self-report or partner report | Yes (*) | Restricts to or adjusts for 1 key variables (*) | Regular follow-up (*) | Mean or median follow-up of at least 3 months (*) | Subjects lost to follow up unlikely to introduce bias (*) | **6** |
| Partridge 2007 | Somewhat representative (*) | Male university students | Drawn from the same community as exposed cohort (*) | Clinician assessment (*) | Yes (*) | Restricts to or adjusts for 1 key variables (*) | Regular follow-up (*) | Mean or median follow-up of at least 3 months (*) | Subjects lost to follow up unlikely to introduce bias (*) | **7** |
| Shapiro 2022 | Somewhat representative (*) | Female university students and their male partners | Drawn from the same community as exposed cohort (*) | Clinician assessment (*) | Yes (*) | Restricts to or adjusts for at least 2 key variables (**) | Regular follow-up (*) | Mean or median follow-up of less than 3 months | Not reported | **7** |
| VanBuskirk 2011 | Somewhat representative (*) | Male university students | Drawn from the same community as exposed cohort (*) | Clinician assessment (*) | Yes (*) | Restricts to or adjusts for 1 key variables (*) | Regular follow-up (*) | Mean or median follow-up of at least 3 months (*) | Subjects lost to follow up unlikely to introduce bias (*) | **7** |

**Supplementary Table 9: Risk of bias assessment, randomized trials**

| | Randomization | | | | Assignment to intervention | | | | |
|---|---|---|---|---|---|---|---|---|---|
| **First author & year** | **Was the allocation sequence random?** | **Was the allocation sequence concealed until participants were enrolled and assigned to interventions?** | **Did baseline differences between intervention groups suggest a problem with the randomization process?** | **Risk-of-bias judgement** | **Were participants aware of their assigned intervention during the trial?** | **Were carers and people delivering the interventions aware of participants' assigned intervention during the trial?** | **Were there deviations from the intended intervention that arose because of the trial context?** | **Was an appropriate analysis used to estimate the effect of assignment to intervention?** | **Risk-of-bias judgement** |
| Gray 2010 | Yes | Yes | No | **Low** | Yes | Yes | No | Yes | **Low** |
| Smith 2021 | Yes | Yes | No | **Low** | Yes | Yes | No | Yes | **Low** |
| Tobian 2009 | Yes | Yes | No | **Low** | Yes | Yes | No | Yes | **Low** |
| Tobian 2012 | Yes | Yes | Probably no | **Low** | Yes | Yes | No | Probably yes | **Low** |
| Wawer 2011 | Yes | Yes | No | **Low** | Yes | Yes | No | Yes | **Low** |

| | Adherence to intervention | | | | | | Missing outcome data | | |
|---|---|---|---|---|---|---|---|---|---|
| **First author & year** | **Were participants aware of their assigned intervention during the trial?** | **Were carers and people delivering the interventions aware of participants' assigned intervention during the trial?** | **Were important non-protocol interventions balanced across intervention groups?** | **Were there failures in implementing the intervention that could have affected the outcome?** | **Was there non-adherence to the assigned intervention regimen that could have affected participants' outcomes?** | **Risk-of-bias judgement** | **Were data for this outcome available for all, or nearly all, participants randomized?** | **Is there evidence that the result was not biased by missing outcome data?`** | **Risk-of-bias judgement** |
| Gray 2010 | Yes | Yes | Probably yes | No | No | **Low** | No | Yes | **Low** |
| Smith 2021 | Yes | Yes | Yes | No | No | **Low** | No | Probably no | **Some concern** |
| Tobian 2009 | Yes | Yes | Probably yes | No | No | **Low** | No | Yes | **Low** |
| Tobian 2012 | Yes | Yes | Probably yes | No | No | **Low** | No | Probably no | **Some concern** |
| Wawer 2011 | Yes | Yes | Probably yes | No | No | **Low** | Yes | Probably yes | **Low** |

| | Outcome measurement | | | | | Selection of reported result | | | |
|---|---|---|---|---|---|---|---|---|---|
| **First author & year** | **Was the method of measuring the outcome inappropriate?** | **Could measurement or ascertainment of the outcome have differed between intervention groups?** | **Were outcome assessors aware of the intervention received by study participants?** | **Could assessment of the outcome have been influenced by knowledge of intervention received?** | **Risk-of-bias judgement** | **Were the data that produced this result analyzed in accordance with a pre-specified analysis plan that was finalized before unblinded outcome data were available for analysis?** | **Is the numerical result being assessed likely to have been selected, on the basis of the results, from multiple eligible outcome measurements within the outcome domain?** | **Is the numerical result being assessed likely to have been selected, on the basis of the results, from multiple eligible analyses of the data?** | **Risk-of-bias judgement** |
| Gray 2010 | No | Probably no | No | No | **Some concern** | Probably no | No | No | **Low** |
| Smith 2021 | No | Probably no | No | No | **Some concern** | Probably no | No | No | **Low** |

| | | | | | | | | |
|---|---|---|---|---|---|---|---|---|
| Tobian 2009 | No | Probably no | No | No | **Some concern** | Probably yes | No | No | **Low** |
| Tobian 2012 | No | Probably no | No | No | **Some concern** | Probably no | No | No | **Low** |
| Wawer 2011 | No | No | No | No | **Low** | Probably yes | No | No | **Low** |

**Supplementary Figure 1: Meta-analysis of studies of male circumcision and high-risk HPV prevalence in males**

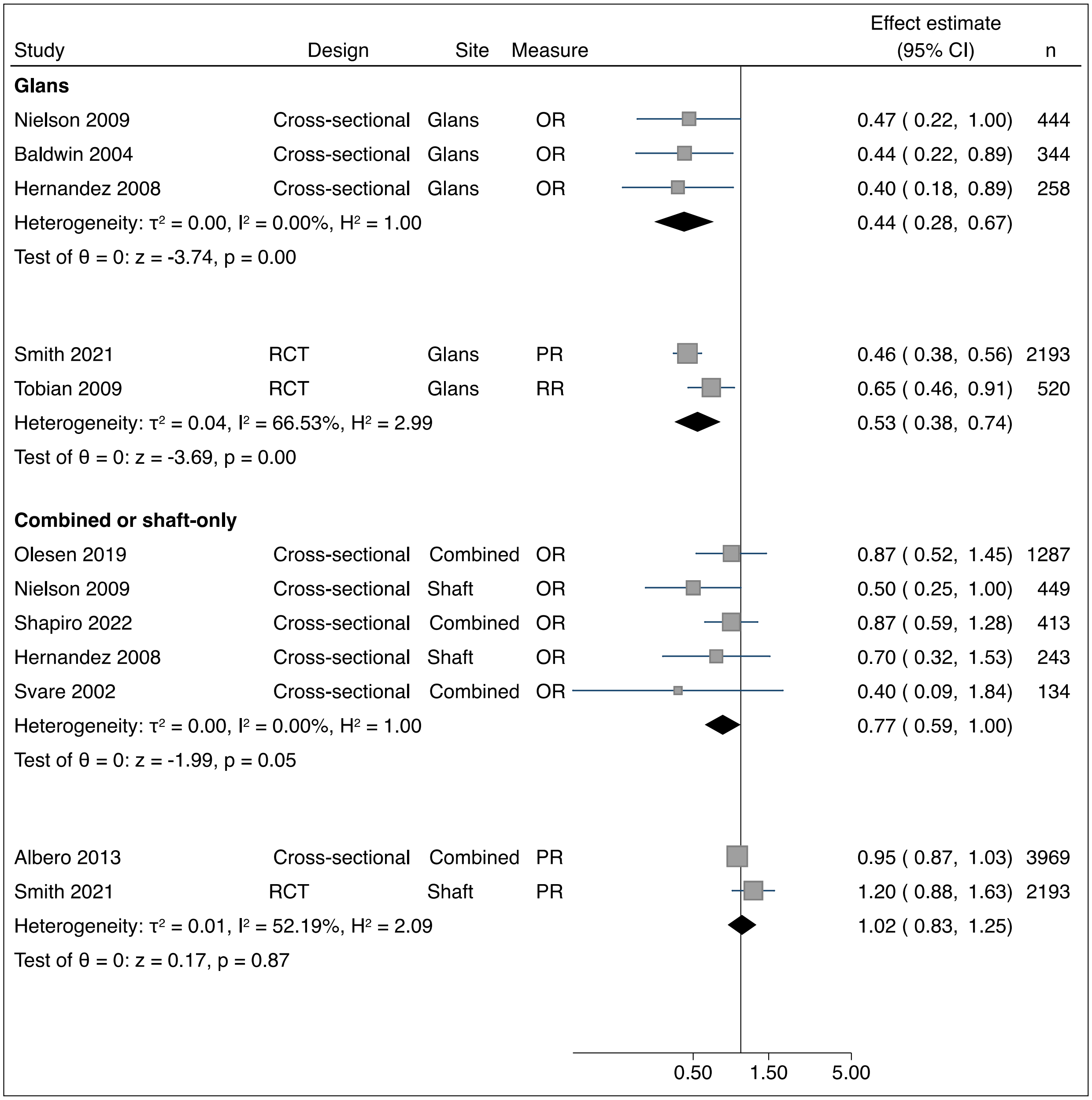

Abbreviations: HPV, human papillomavirus; OR, odds ratio; PR, prevalence ratio; RCT, randomized controlled trial; RR, risk ratio

**Supplementary Figure 2: Meta-analysis of studies of male circumcision and low-risk HPV prevalence in males**

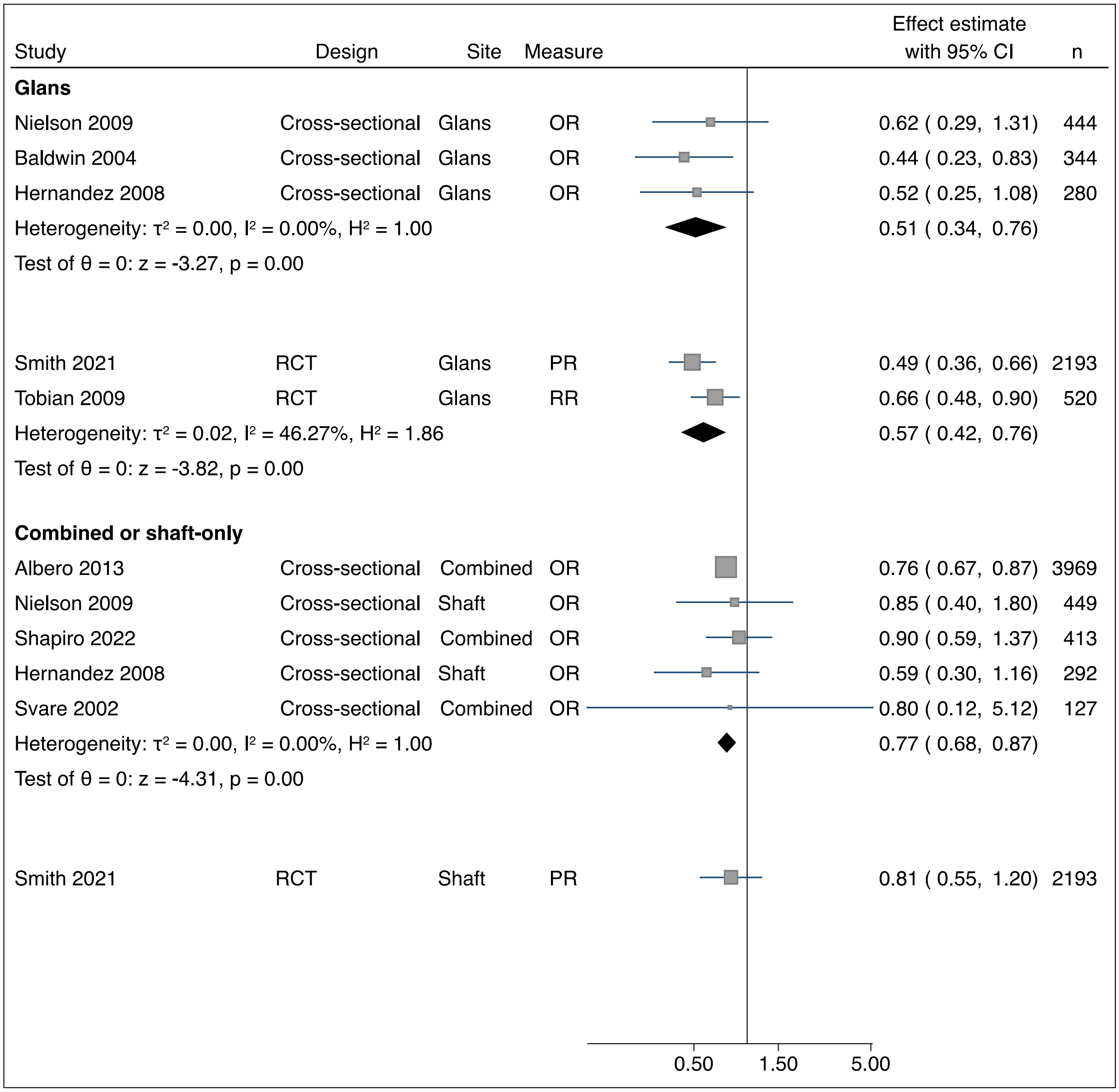

Abbreviations: HPV, human papillomavirus; OR, odds ratio; PR, prevalence ratio; RCT, randomized controlled trial; RR, risk ratio

**Supplementary Figure 3: Meta-analysis of studies of male circumcision and high-risk HPV incidence in males**

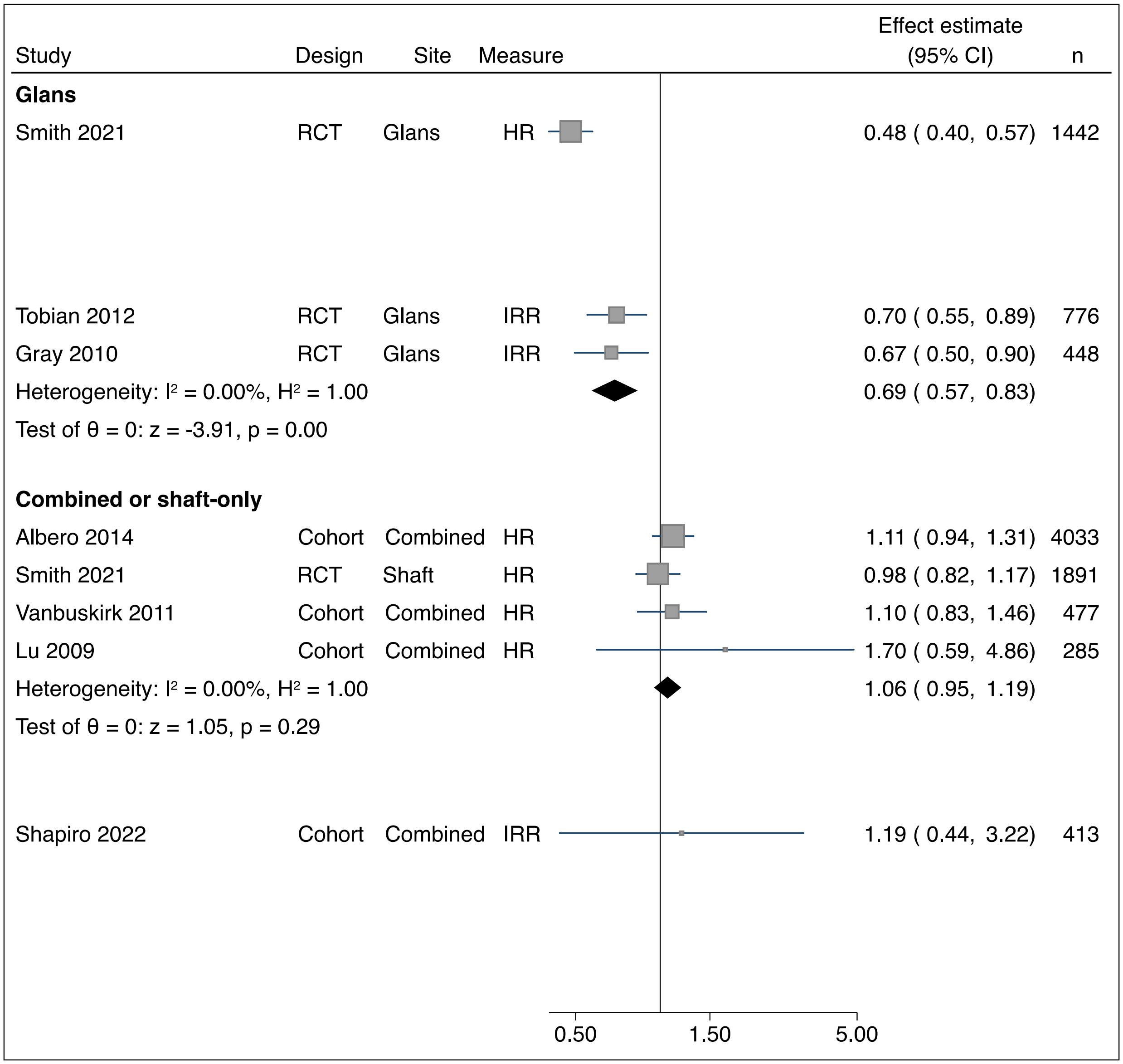

Abbreviations: HPV, human papillomavirus; HR, hazard ratio; IRR, incidence rate ratio; RCT, randomized controlled trial

**Supplementary Figure 4: Meta-analysis of studies of male circumcision and low-risk HPV incidence in males**

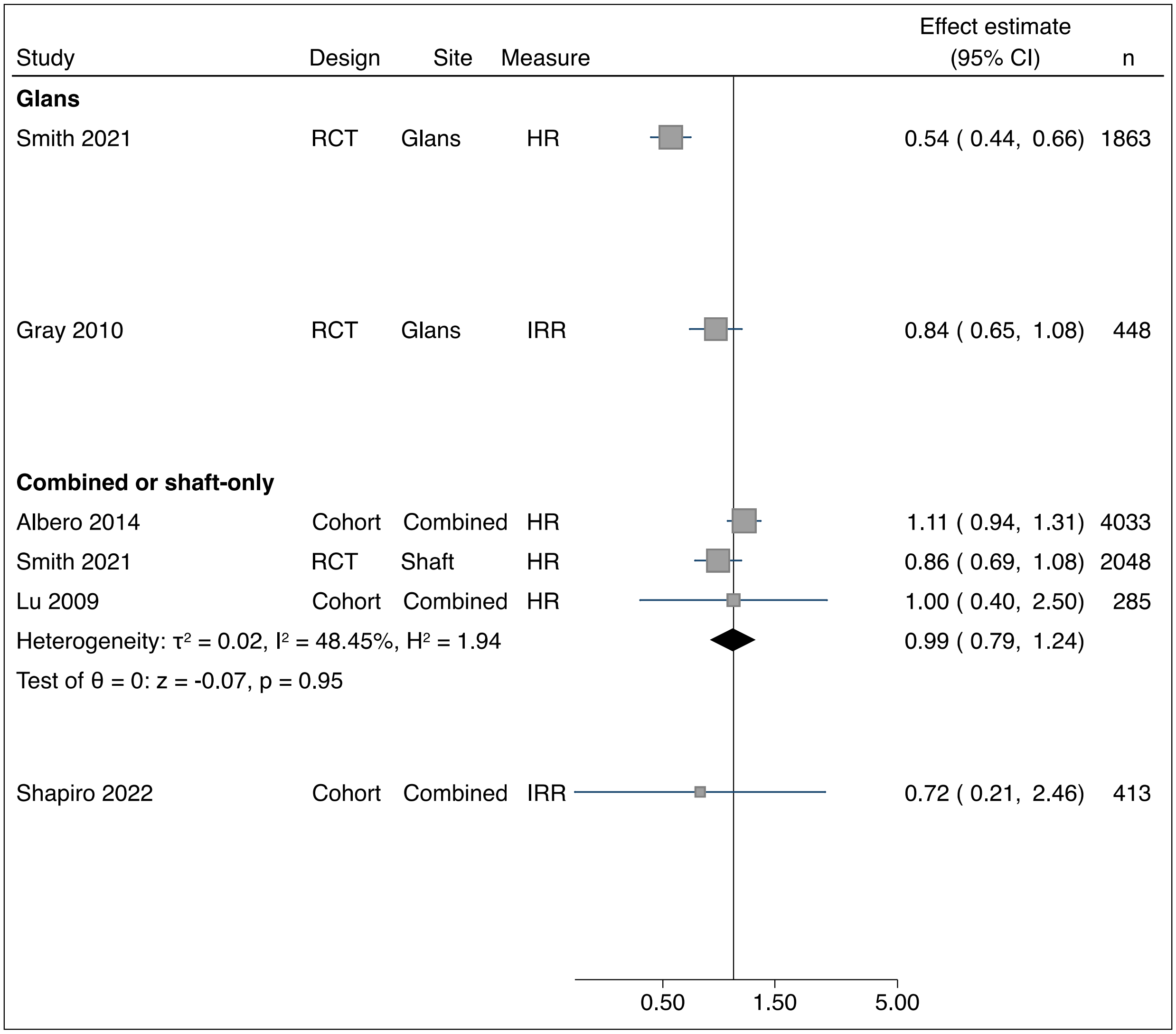

Abbreviations: HPV, human papillomavirus; HR, hazard ratio; IRR, incidence rate ratio; RCT, randomized controlled trial

**Supplementary Figure 5: Meta-analysis of studies of male circumcision and high-risk HPV clearance in males**

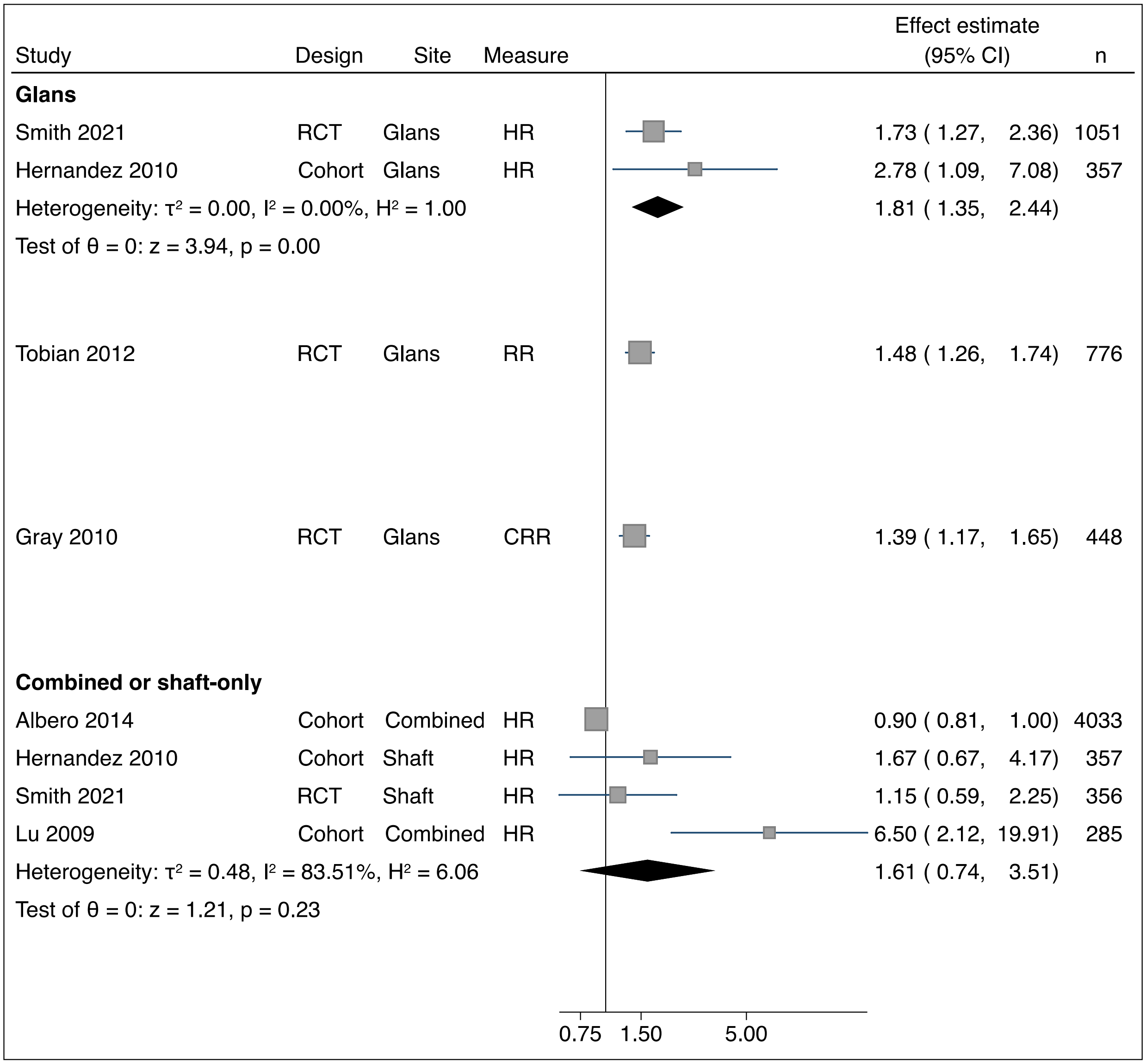

Abbreviations: CRR, clearance rate ratio; HPV, human papillomavirus; HR, hazard ratio; RCT, randomized controlled trial; RR, risk ratio

**Supplementary Figure 6: Meta-analysis of studies of male circumcision and low-risk HPV clearance in males**

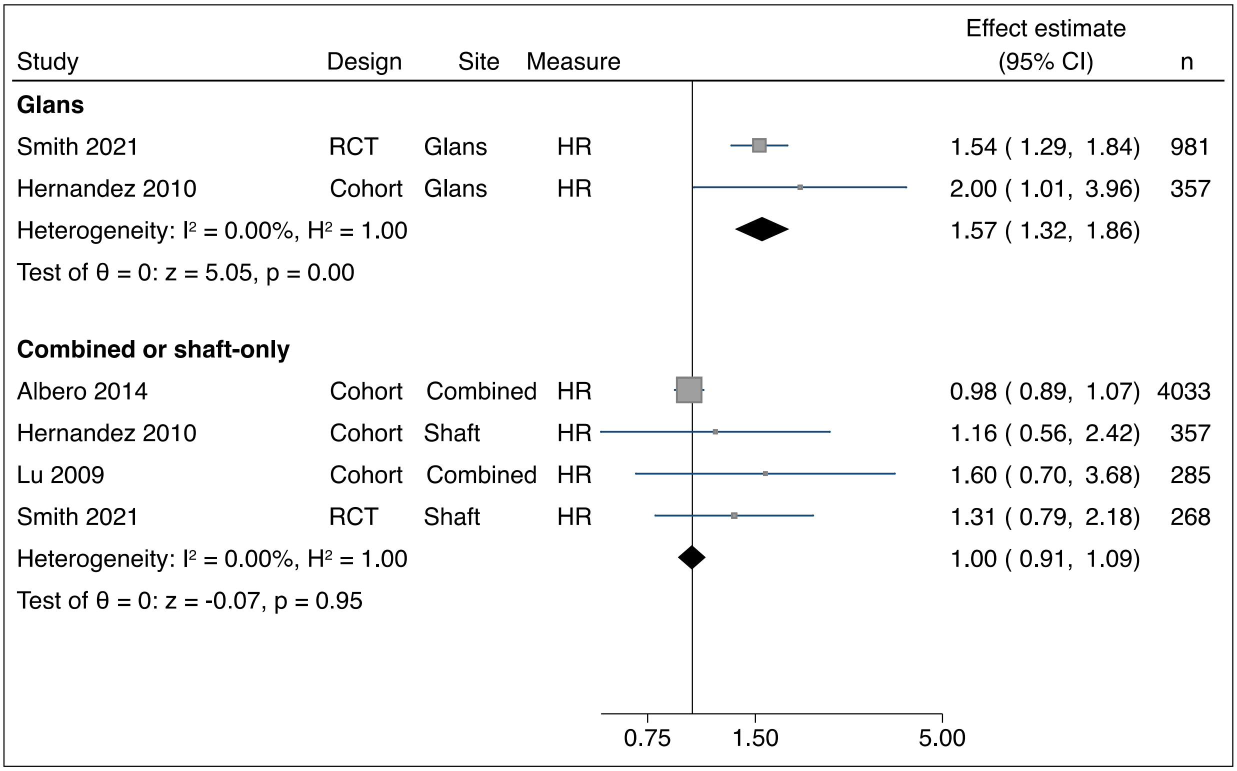

Abbreviations: CRR, clearance rate ratio; HPV, human papillomavirus; HR, hazard ratio; RCT, randomized controlled trial

**Supplementary Figure 7: Meta-analysis of studies of male circumcision and HPV prevalence in males, low risk of bias studies**

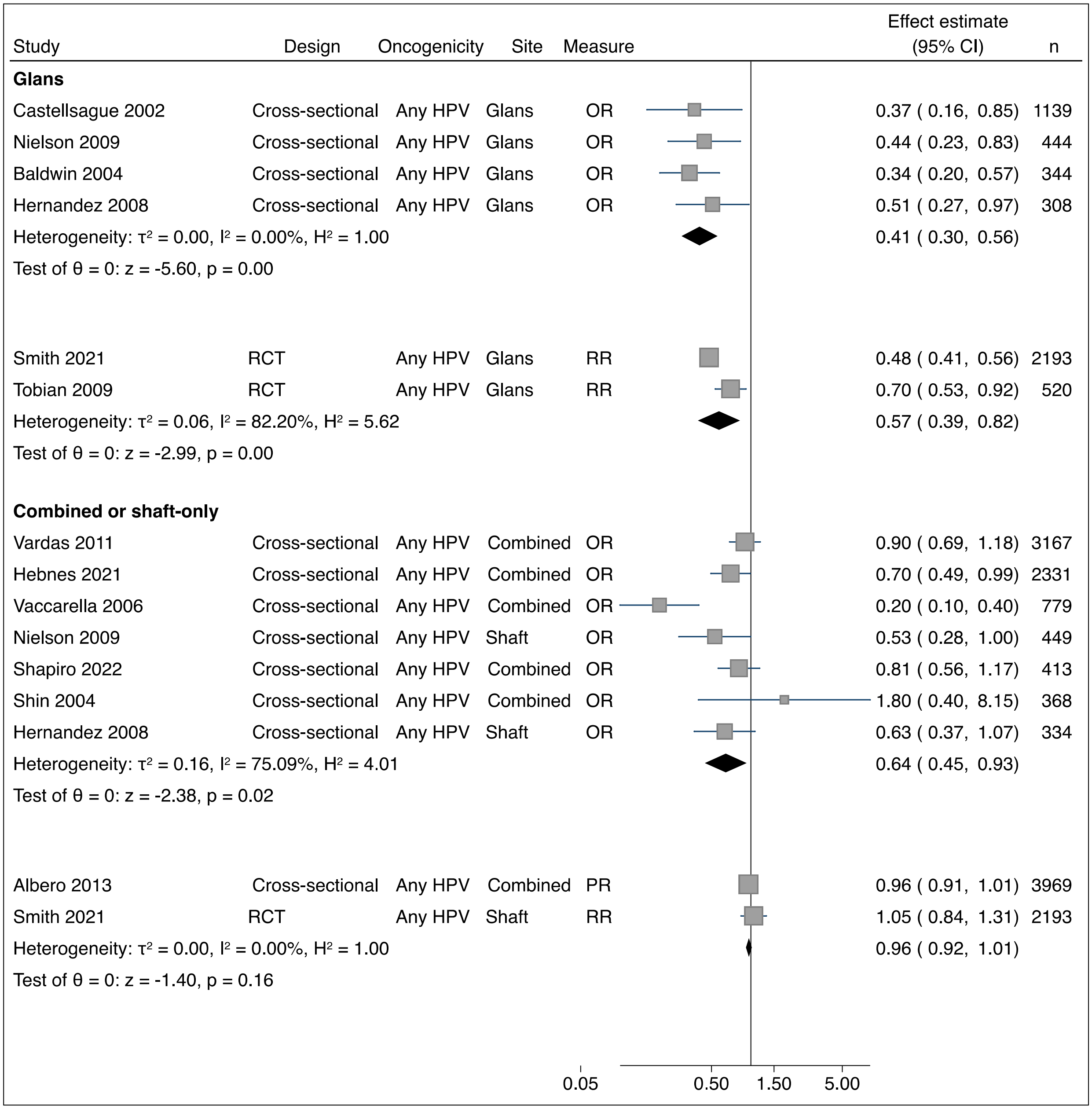

Abbreviations: HPV, human papillomavirus; OR, odds ratio; PR, prevalence ratio; RCT, randomized controlled trial; RR, risk ratio

**Supplementary Figure 8: Meta-analysis of studies of male circumcision and HPV incidence in males, low risk of bias studies**

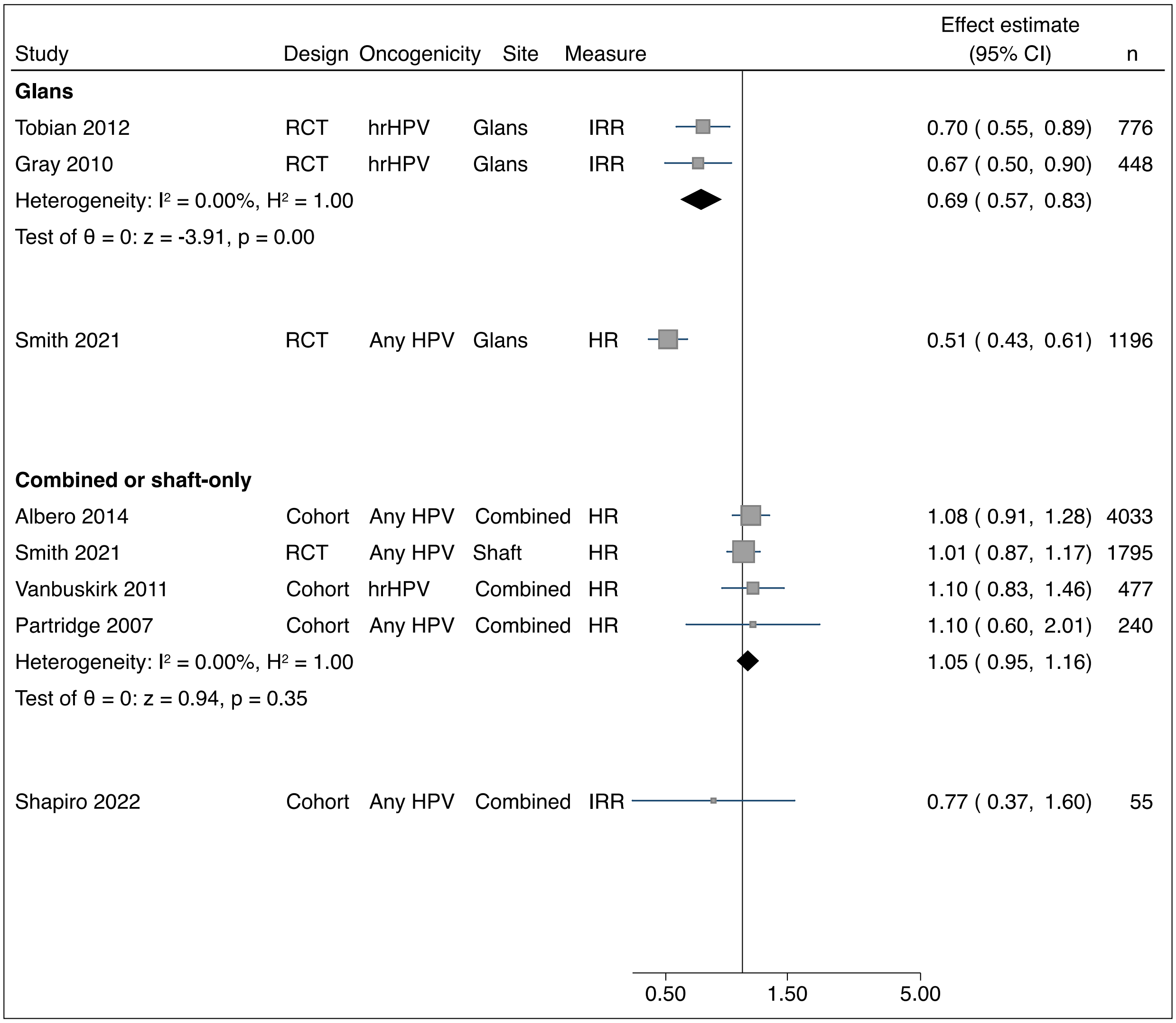

Abbreviations: HPV, human papillomavirus; HR, hazard ratio; hrHPV, high-risk HPV; IRR, incidence rate ratio; RCT, randomized controlled trial

**Supplementary Figure 9: Meta-analysis of studies of male circumcision and HPV clearance in males, low risk of bias studies**

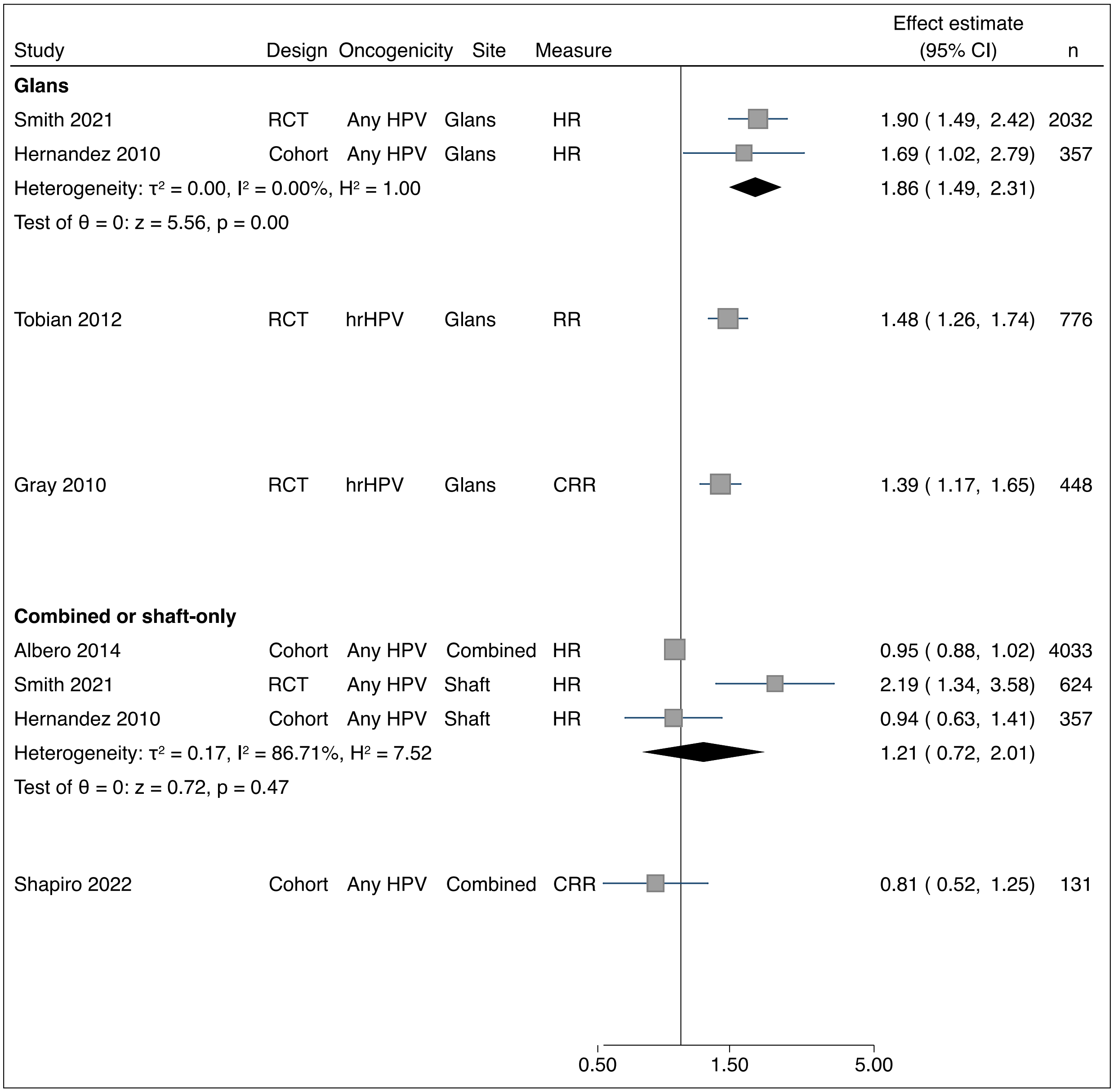

Abbreviations: CRR, clearance rate ratio; HPV, human papillomavirus; HR, hazard ratio; hrHPV, high-risk HPV; RCT, randomized controlled trial; RR, risk ratio

**Supplementary Figure 10: Meta-analysis of studies of male circumcision and HPV prevalence in males, by control for confounding**

| Study | Design | Oncogenicity | Site | Measure | Effect estimate (95% CI) | n |
|---|---|---|---|---|---|---|
| **Glans, control for confounding** | | | | | | |
| Castellsague 2002 | Cross-sectional | Any HPV | Glans | OR | 0.37 ( 0.16, 0.85) | 1139 |
| Nielson 2009 | Cross-sectional | Any HPV | Glans | OR | 0.44 ( 0.23, 0.83) | 444 |
| Baldwin 2004 | Cross-sectional | Any HPV | Glans | OR | 0.34 ( 0.20, 0.57) | 344 |
| Hernandez 2008 | Cohort | Any HPV | Glans | OR | 0.51 ( 0.27, 0.97) | 308 |
| Heterogeneity: $\tau^2$ = 0.00, $I^2$ = 0.00%, $H^2$ = 1.00 | | | | | 0.41 ( 0.30, 0.56) | |
| Test of θ = 0: z = -5.60, p = 0.00 | | | | | | |
| Smith 2021 | RCT | Any HPV | Glans | RR | 0.48 ( 0.41, 0.56) | 2193 |
| Tobian 2009 | RCT | Any HPV | Glans | RR | 0.70 ( 0.53, 0.92) | 520 |
| Heterogeneity: $\tau^2$ = 0.06, $I^2$ = 82.20%, $H^2$ = 5.62 | | | | | 0.57 ( 0.39, 0.82) | |
| Test of θ = 0: z = -2.99, p = 0.00 | | | | | | |
| **Glans, no control for confounding** | | | | | | |
| Bleeker 2005 | Cross-sectional | Any HPV | Glans | OR | 0.98 ( 0.41, 2.35) | 253 |
| Da Rocha 2015 | Cross-sectional | Any HPV | Glans | OR | 0.52 ( 0.04, 7.03) | 182 |
| Rocha 2012 | Cross-sectional | Any HPV | Glans | OR | 1.32 ( 0.06, 28.87) | 37 |
| Heterogeneity: $\tau^2$ = 0.00, $I^2$ = 0.00%, $H^2$ = 1.00 | | | | | 0.94 ( 0.42, 2.10) | |
| Test of θ = 0: z = -0.15, p = 0.88 | | | | | | |
| **Combined or shaft-only, control for confounding** | | | | | | |
| Vardas 2011 | Cross-sectional | Any HPV | Combined | OR | 0.90 ( 0.69, 1.18) | 3167 |
| Hebnes 2021 | Cross-sectional | Any HPV | Combined | OR | 0.70 ( 0.49, 0.99) | 2331 |
| Lajous 2005 | Cohort | Any HPV | Combined | OR | 0.48 ( 0.30, 0.77) | 925 |
| Vaccarella 2006 | Cross-sectional | Any HPV | Combined | OR | 0.20 ( 0.10, 0.40) | 779 |
| Nielson 2009 | Cross-sectional | Any HPV | Shaft | OR | 0.53 ( 0.28, 1.00) | 449 |
| Shapiro 2022 | Cohort | Any HPV | Combined | OR | 0.81 ( 0.56, 1.17) | 413 |
| Shin 2004 | Cross-sectional | Any HPV | Combined | OR | 1.80 ( 0.40, 8.15) | 368 |
| Hernandez 2008 | Cohort | Any HPV | Shaft | OR | 0.63 ( 0.37, 1.07) | 334 |
| Svare 2002 | Cross-sectional | Any HPV | Combined | OR | 0.20 ( 0.06, 0.63) | 198 |
| Heterogeneity: $\tau^2$ = 0.17, $I^2$ = 73.75%, $H^2$ = 3.81 | | | | | 0.58 ( 0.41, 0.81) | |
| Test of θ = 0: z = -3.21, p = 0.00 | | | | | | |
| Albero 2013 | Cohort | Any HPV | Combined | PR | 0.96 ( 0.91, 1.01) | 3969 |
| Smith 2021 | RCT | Any HPV | Shaft | RR | 1.05 ( 0.84, 1.31) | 2193 |
| Heterogeneity: $\tau^2$ = 0.00, $I^2$ = 0.00%, $H^2$ = 1.00 | | | | | 0.96 ( 0.92, 1.01) | |
| Test of θ = 0: z = -1.40, p = 0.16 | | | | | | |
| **Combined or shaft-only, no control for confounding** | | | | | | |
| Olesen 2019 | Cross-sectional | hrHPV | Combined | OR | 0.87 ( 0.52, 1.45) | 1287 |
| Mbulawa 2009 | Cross-sectional | Any HPV | Combined | OR | 0.54 ( 0.20, 1.42) | 298 |
| Ogilvie 2009 | Cross-sectional | Any HPV | Combined | OR | 1.14 ( 0.67, 1.94) | 262 |
| Rombaldi 2006 | Cross-sectional | Any HPV | Combined | OR | 2.07 ( 0.46, 9.35) | 99 |
| Heterogeneity: $\tau^2$ = 0.00, $I^2$ = 0.00%, $H^2$ = 1.00 | | | | | 0.96 ( 0.68, 1.34) | |
| Test of θ = 0: z = -0.27, p = 0.79 | | | | | | |

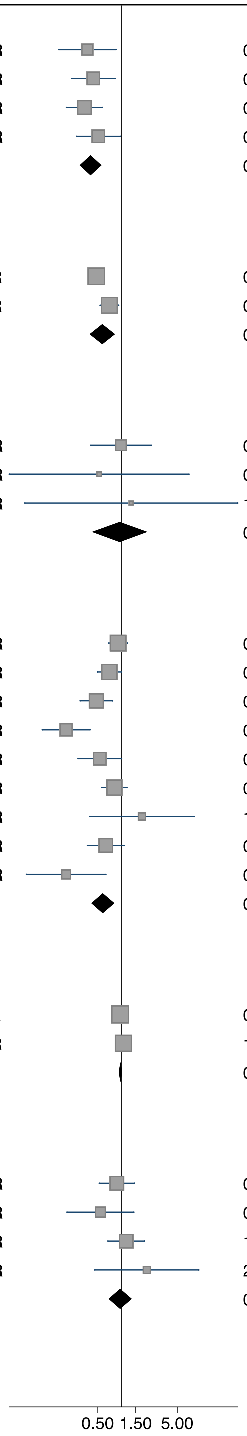

Abbreviations: HPV, human papillomavirus; hrHPV, high-risk HPV; OR, odds ratio; PR, prevalence ratio; RR, risk ratio

**Supplementary Figure 11: Leave-one-out analysis for studies of male circumcision and HPV prevalence in males**

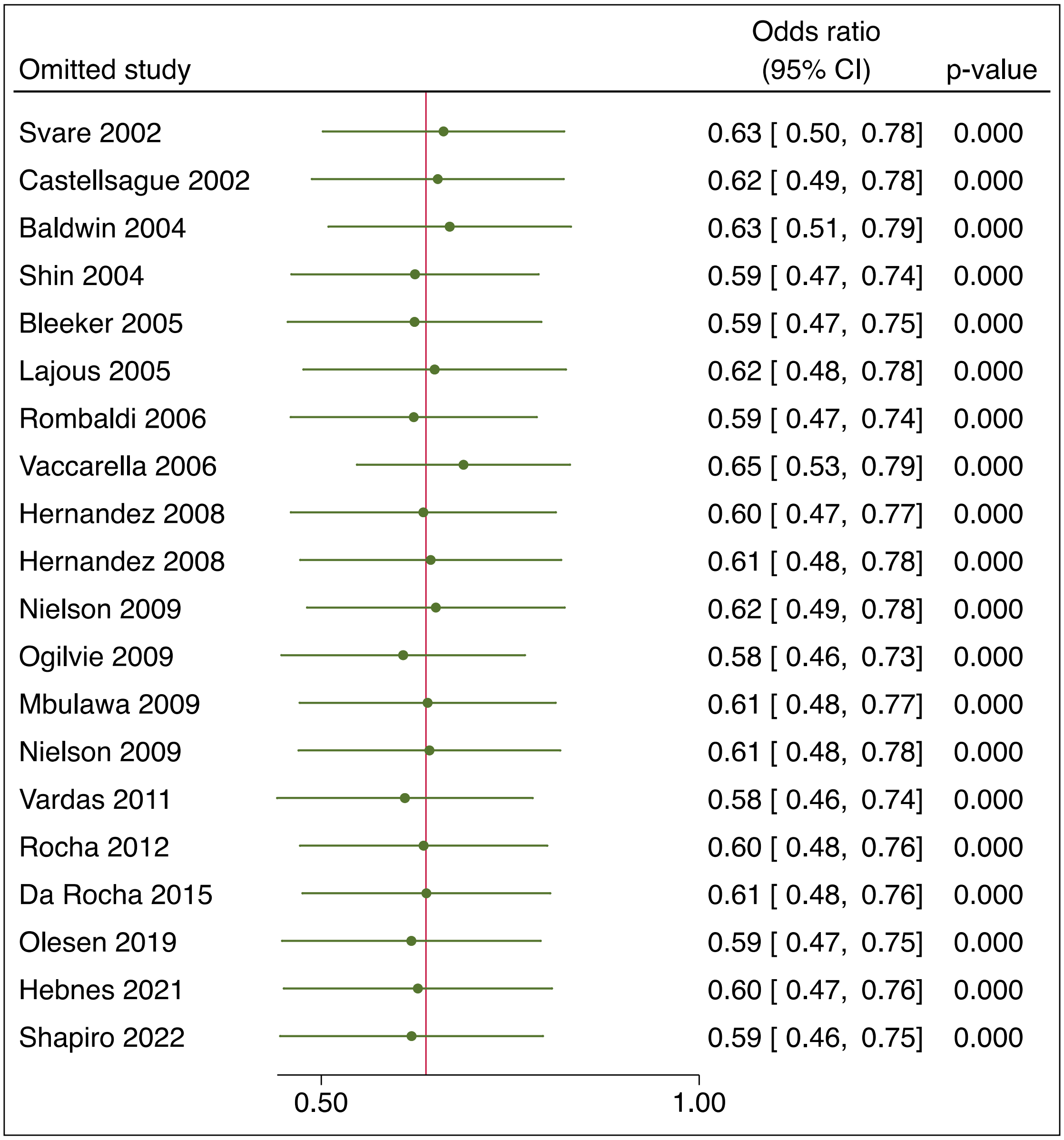

Abbreviations: CI, confidence interval; HPV, human papillomavirus; REML, restricted maximum likelihood

**Supplementary Figure 12: Funnel plot for studies of male circumcision and HPV prevalence in males**

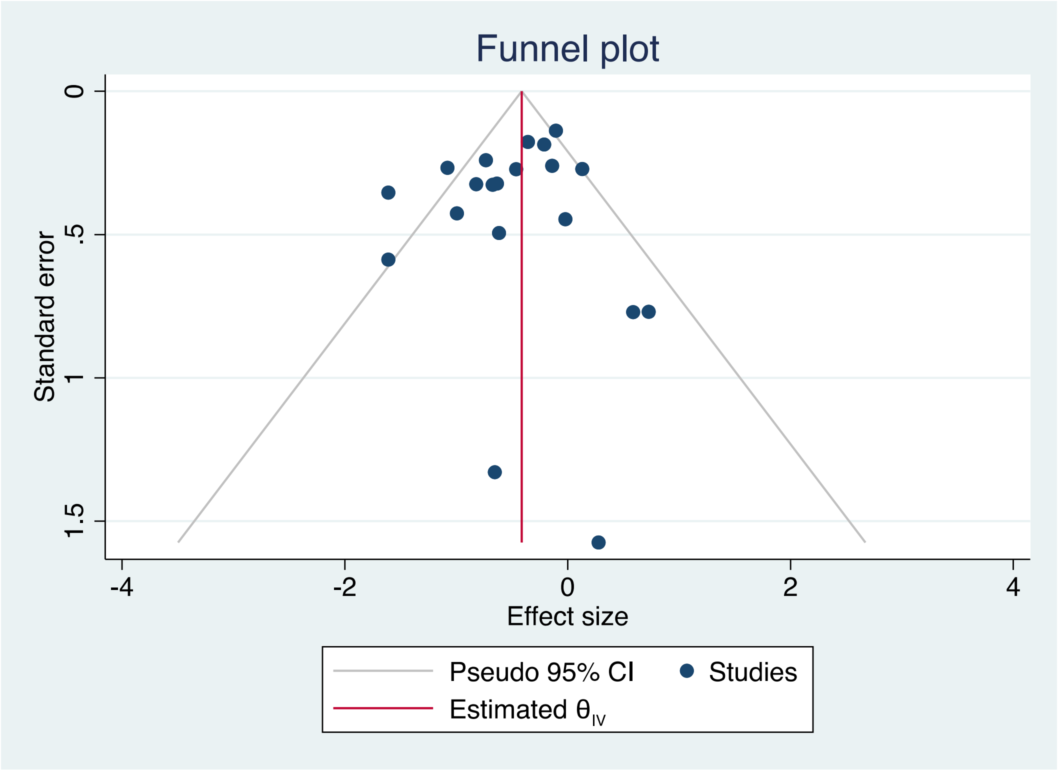

Abbreviations: CI, confidence interval; HPV, human papillomavirus

### PRISMA 2020 Checklist

| Section and Topic | Item # | Checklist item | Page where item is reported |
|---|---|---|---|
| **TITLE** | | | |
| Title | 1 | Identify the report as a systematic review. | 1 |
| **ABSTRACT** | | | |
| Abstract | 2 | See the PRISMA 2020 for Abstracts checklist. | 2 |
| **INTRODUCTION** | | | |
| Rationale | 3 | Describe the rationale for the review in the context of existing knowledge. | 5 |
| Objectives | 4 | Provide an explicit statement of the objective(s) or question(s) the review addresses. | 5 |
| **METHODS** | | | |
| Eligibility criteria | 5 | Specify the inclusion and exclusion criteria for the review and how studies were grouped for the syntheses. | 5-6 |
| Information sources | 6 | Specify all databases, registers, websites, organisations, reference lists and other sources searched or consulted to identify studies. Specify the date when each source was last searched or consulted. | 6 |
| Search strategy | 7 | Present the full search strategies for all databases, registers and websites, including any filters and limits used. | S2 |
| Selection process | 8 | Specify the methods used to decide whether a study met the inclusion criteria of the review, including how many reviewers screened each record and each report retrieved, whether they worked independently, and if applicable, details of automation tools used in the process. | 6 |
| Data collection process | 9 | Specify the methods used to collect data from reports, including how many reviewers collected data from each report, whether they worked independently, any processes for obtaining or confirming data from study investigators, and if applicable, details of automation tools used in the process. | 6 |
| Data items | 10a | List and define all outcomes for which data were sought. Specify whether all results that were compatible with each outcome domain in each study were sought (e.g. for all measures, time points, analyses), and if not, the methods used to decide which results to collect. | 6-7 |
| | 10b | List and define all other variables for which data were sought (e.g. participant and intervention characteristics, funding sources). Describe any assumptions made about any missing or unclear information. | 6-7 |
| Study risk of bias assessment | 11 | Specify the methods used to assess risk of bias in the included studies, including details of the tool(s) used, how many reviewers assessed each study and whether they worked independently, and if applicable, details of automation tools used in the process. | 7 |
| Effect measures | 12 | Specify for each outcome the effect measure(s) (e.g. risk ratio, mean difference) used in the synthesis or presentation of results. | 7 |
| Synthesis methods | 13a | Describe the processes used to decide which studies were eligible for each synthesis (e.g. tabulating the study intervention characteristics and comparing against the planned groups for each synthesis (item #5)). | 7 |
| | 13b | Describe any methods required to prepare the data for presentation or synthesis, such as handling of missing summary statistics, or data conversions. | 7 |
| | 13c | Describe any methods used to tabulate or visually display results of individual studies and syntheses. | 7 |
| | 13d | Describe any methods used to synthesize results and provide a rationale for the choice(s). If meta-analysis was performed, describe the model(s), method(s) to identify the presence and extent of statistical heterogeneity, and software package(s) used. | 7 |
| | 13e | Describe any methods used to explore possible causes of heterogeneity among study results (e.g. subgroup analysis, meta-regression). | 7 |
| | 13f | Describe any sensitivity analyses conducted to assess robustness of the synthesized results. | 7 |
| Reporting bias assessment | 14 | Describe any methods used to assess risk of bias due to missing results in a synthesis (arising from reporting biases). | 7 |
| Certainty assessment | 15 | Describe any methods used to assess certainty (or confidence) in the body of evidence for an outcome. | 7 |

### PRISMA 2020 Checklist

| Section and Topic | Item # | Checklist item | Page where item is reported |
|---|---|---|---|
| **RESULTS** | | | |
| Study selection | 16a | Describe the results of the search and selection process, from the number of records identified in the search to the number of studies included in the review, ideally using a flow diagram. | 8 |
| | 16b | Cite studies that might appear to meet the inclusion criteria, but which were excluded, and explain why they were excluded. | S3-5 |
| Study characteristics | 17 | Cite each included study and present its characteristics. | 9-10, 24-27, S6-23 |
| Risk of bias in studies | 18 | Present assessments of risk of bias for each included study. | 11, S24-29 |
| Results of individual studies | 19 | For all outcomes, present, for each study: (a) summary statistics for each group (where appropriate) and (b) an effect estimate and its precision (e.g. confidence/credible interval), ideally using structured tables or plots. | 10-11, 30-33, S30-35 |
| Results of syntheses | 20a | For each synthesis, briefly summarise the characteristics and risk of bias among contributing studies. | 11-12, S36-38 |
| | 20b | Present results of all statistical syntheses conducted. If meta-analysis was done, present for each the summary estimate and its precision (e.g. confidence/credible interval) and measures of statistical heterogeneity. If comparing groups, describe the direction of the effect. | 11-12, 30-33, S30-35 |
| | 20c | Present results of all investigations of possible causes of heterogeneity among study results. | 11-12, 28 |
| | 20d | Present results of all sensitivity analyses conducted to assess the robustness of the synthesized results. | 11-12, S36-40 |
| Reporting biases | 21 | Present assessments of risk of bias due to missing results (arising from reporting biases) for each synthesis assessed. | NA |
| Certainty of evidence | 22 | Present assessments of certainty (or confidence) in the body of evidence for each outcome assessed. | 10-11, 30-33, S30-35 |
| **DISCUSSION** | | | |
| Discussion | 23a | Provide a general interpretation of the results in the context of other evidence. | 12 |
| | 23b | Discuss any limitations of the evidence included in the review. | 14 |
| | 23c | Discuss any limitations of the review processes used. | 14 |
| | 23d | Discuss implications of the results for practice, policy, and future research. | 14 |
| **OTHER INFORMATION** | | | |
| Registration and protocol | 24a | Provide registration information for the review, including register name and registration number, or state that the review was not registered. | 8 |
| | 24b | Indicate where the review protocol can be accessed, or state that a protocol was not prepared. | 8 |
| | 24c | Describe and explain any amendments to information provided at registration or in the protocol. | 8 |
| Support | 25 | Describe sources of financial or non-financial support for the review, and the role of the funders or sponsors in the review. | 8 |
| Competing interests | 26 | Declare any competing interests of review authors. | 15 |
| Availability of data, code and other materials | 27 | Report which of the following are publicly available and where they can be found: template data collection forms; data extracted from included studies; data used for all analyses; analytic code; any other materials used in the review. | 15 |

*From:* Page MJ, McKenzie JE, Bossuyt PM, Boutron I, Hoffmann TC, Mulrow CD, et al. The PRISMA 2020 statement: an updated guideline for reporting systematic reviews. BMJ 2021;372:n71. doi:
